# Reduced Penetrance is Common Among Predicted Loss-of-Function Variants and is Likely Driven by Residual Allelic Activity

**DOI:** 10.1101/2024.09.23.24314008

**Authors:** David R. Blair, Neil Risch

## Abstract

Loss-of-function genetic variants (LoFs) often result in severe phenotypes, including autosomal dominant diseases driven by haploinsufficiency. Due to low carrier frequencies, their penetrance is generally unknown but typically variable. Here, we investigate the penetrance of >6,000 predicted LoFs (pLoFs) linked to 91 haploinsufficient diseases using a cohort of ≈24,000 carriers with linked electronic health record data. We find evidence for widespread reduced penetrance, which persisted after accounting for variant annotation artifacts, missed diagnoses, and incomplete clinical data. We thus hypothesized that many pLoFs have incomplete penetrance, which may be driven by residual allelic activity. To test this, we trained machine learning models to predict pLoF penetrance using variant-specific genomic features that may correlate with incomplete loss-of-function. The models were predictive of pLoF penetrance across a range of diseases and variant types, including those with prior clinical evidence for pathogenicity. This suggests that many pLoFs have incomplete penetrance due to residual allelic activity, complicating disease prognostication in asymptomatic carriers.

## Introduction

Exome (ES) and genome sequencing (GS) are now first-tier tests for rare disease diagnosis^1–6^. Given this success, there is growing interest in applying these technologies to asymptomatic individuals^7–17^, including those incidentally found to carry pathogenic variants during routine diagnostic testing^18^. The utility of sequencing for prognosis remains uncertain^19–21^. Generally, a test’s utility for prognosis is quantified using its positive predictive value. For genetic testing, this statistic is driven by both the accuracy of the genotype calls and their penetrance, where penetrance is defined as the (age-dependent) probability that a carrier will manifest disease. Although imperfect, genotype calling accuracy is relatively high^22,23^. Alternatively, penetrance for most genotypes is unknown. It can range from 0 (no associated disease risk) to 1 (certain disease manifestation), vary with age, and be modified by additional factors, including polygenic background^24–26^ and environmental exposures^27^. For diagnostic applications, accurate penetrance estimates are not critical. Patients already express a disease phenotype, so laboratories must only determine if variants are causal (i.e. pathogenic) or benign^28^. Variant interpretation in asymptomatic cases is more complex. Laboratories and clinicians should be able to express how likely the variants are to cause disease in the future. This risk is, of course, inextricably tied to penetrance.

Penetrance is notoriously difficult to estimate^29^. For a few pathogenic genotypes that are unusually common (typically due to founder events), accurate penetrance estimation is possible^30–33^. Generally, however, pathogenic genotypes are extremely rare. As a result, penetrance is unknown for most. It is tempting to generalize penetrance estimates from a single or few well-characterized variants to others within the same gene, particularly for those variants predicted to have similar molecular impacts (e.g. loss-of-function). However, there is little evidence to support this strong assumption. Recently, population-scale biobanks with linked electronic health record (EHR) and ES or GS data have become widely available^34–41^. These datasets have been used to estimate penetrance using a “genotype-first” approach^42^. Here, pathogenic genotype carriers are identified using the available genetic data, after which their disease expression is measured retrospectively using EHR data. Although imperfect, these analyses suggest widespread incomplete penetrance for pathogenic genotypes^43–45^. This observation has important implications for the interpretation of genetic testing in asymptomatic individuals, as it suggests that the positive predictive value for many genotypes will be unacceptably low. That said, biobanks also have limitations as a resource for estimating penetrance, as fragmented phenotypes and missing clinical data, along with variant annotation artifacts, may lead to deflated estimates^42,46^.

Here, we investigate the range of apparent penetrance for one of the simplest classes of potentially pathogenic mutations: predicted loss-of-function (pLoF) variants in haploinsufficient disease genes. To do so, we uniformly processed the genomic data from two biobanks (the UK Biobank^37^ and the All of Us Research Program^41^; combined *N*>700,000), identifying ≈24,000 pLoF carriers at risk for 91 diseases. On average, biobank pLoFs were associated with disease expression, but consistent with prior studies, penetrance was generally low. We investigated factors that may underly the apparently reduced penetrance, including annotation artifacts, missed diagnoses, and incomplete clinical data. Accounting for these factors increased penetrance, but widespread reduced penetrance persisted. Therefore, we hypothesized that many of these variants may have intrinsically reduced or no penetrance, driven by residual or “leaky” allelic activity. To test this, we trained machine learning models to predict pLoF penetrance using variant-specific genomic features that might correlate with incomplete loss-of-function. These models were able to stratify pLoFs according to their penetrance risk for a range of diseases and variant types, including those previously annotated as pathogenic by diagnostic testing laboratories. This suggests that variants classified as LoF may in fact have varying degrees of residual function^47^, and consequently, their penetrance remains uncertain. Although these analyses were limited to pLoFs in haploinsufficient disease genes, our results likely generalize to other types of disorders (ex: autosomal recessive), variant types (ex: missense) and genetic mechanisms (ex: gain-of-function). Accurately communicating this uncertainty will be crucial for the success of prognostic genomic testing, including population-scale screening programs.

## Results

### Haploinsufficient Disease and pLoF Identification

Using the ClinGen database^48^ (Figure 1A; Step 1), we identified 91 autosomal dominant/pseudo-autosomal dominant Mendelian disorders for which haploinsufficiency is an established mechanism of disease (see Methods). This set of diseases covered a broad range of human pathophysiology, including neurodevelopmental disorders, congenital malformation syndromes, and diseases linked to tumor predisposition. Most (76%) typically present during childhood while the remainder occur throughout adulthood. Figure 1B depicts a summary of the 91 diseases. The individual diseases, along with their annotated information, are provided as Supplementary Dataset 1.

**Figure 1.**
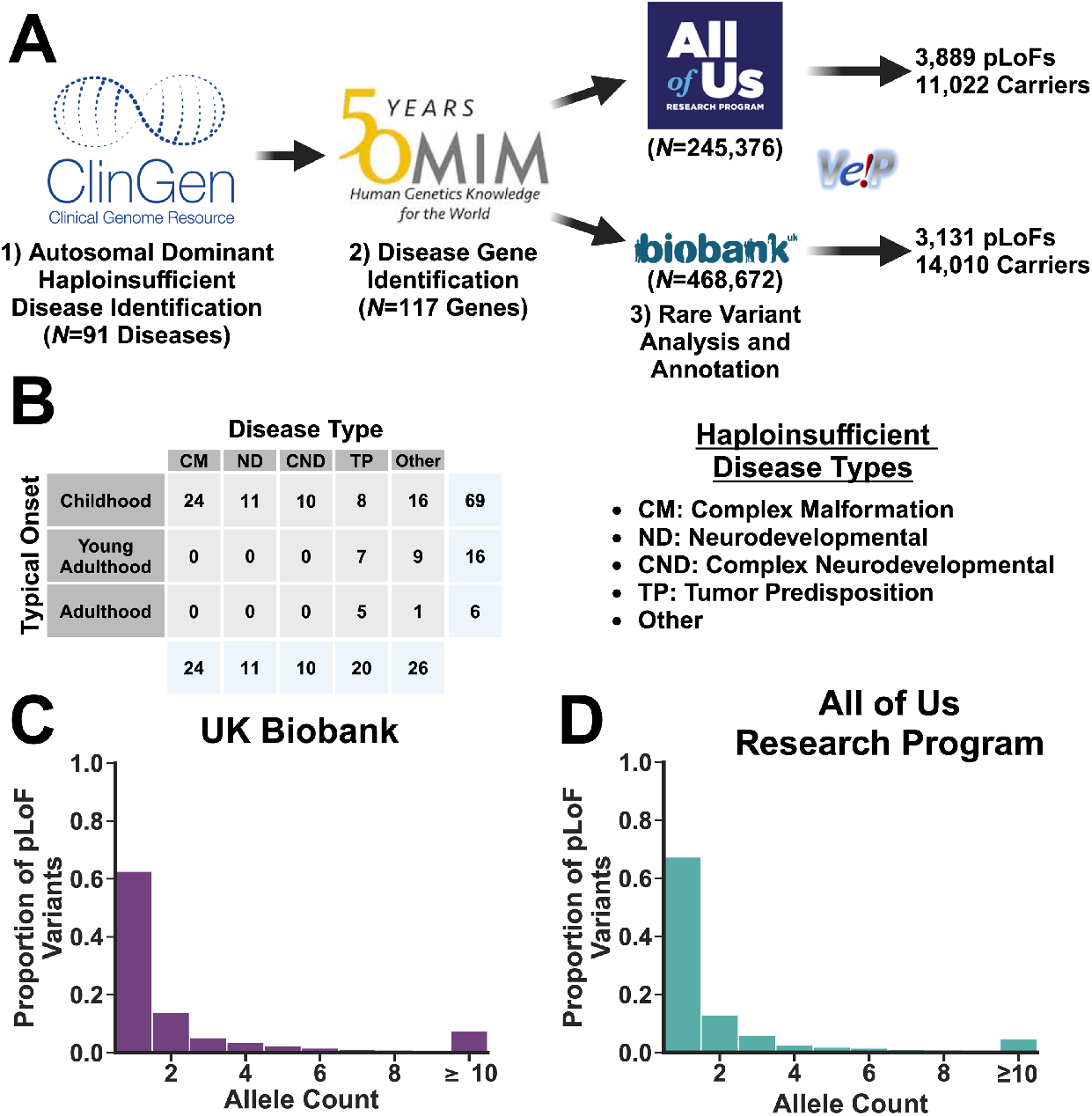
Haploinsufficient Disease and pLoF Identification. (A): Schematic depicting the strategy for haploinsufficient disease identification and biobank sequence processing. (B) Summary of the 91 haploinsufficient diseases investigated in the analysis, stratified by their typical onset class and disease type (left). Disease type definitions are provided to the right. (C, D): The carrier count distributions for the pLoF variants identified in the UK Biobank (C) and the All of Us Research Program (D). Created using Biorender.com.

Following annotation, we linked the diseases to their associated genes using the Online Mendelian Inheritance in Man database^49^ (117 in total; Figure 1A, Step 2). We then systematically identified all rare (biobank-specific carrier frequencies ≤ 0.1%), predicted loss-of-function variants (limited to stop gains, frameshifts and splice changes; pLoFs) within these genes in both biobanks, removing those that likely represent technical artifacts based on sequencing depth and quality scores (Figure 1A, Step 3; see Methods for details). In total, we identified 3,131 unique pLoFs in the UK Biobank (UKBB; total *N*= 468,672) and 3,889 in the All of Us Research Program (AoU; total *N*= 245,376), resulting in a total of 6,247 unique variants (773 occurred in both biobanks).

The individual variants, along with their annotations, are provided as Supplementary Dataset 2 (UKBB) and Supplementary Dataset 3 (AoU). The distributions of pLoF carrier counts for the two biobanks are displayed in Figures 1C and 1D. Most were singletons (63% and 67% in the UKBB and AoU, respectively). Nevertheless, the marginal pLoF carrier frequencies were surprisingly high: 3.0% and 4.5% of subjects in the UKBB and AoU carried at least one pLoF in one of the 117 genes. The substantial difference in carrier frequencies across the two biobanks likely reflects their distinct demographies^37,41^ and ascertainment practices^50,51^. Regardless, these carrier frequencies were much too high for a set of variants that consistently cause disease, suggesting that many had little-to-no phenotypic impact, particularly since they represent only a tiny subset of all possible disease-associated variants.

### pLoFs Have Detectable Phenotypic Effects but with Reduced Penetrance

We restricted our penetrance analyses to pLoFs within genes that were confidently linked to haploinsufficient disorders. Therefore, these variants should have detectable and replicable effects on disease expression. To test this, we constructed a unique comparison group for each disease by identifying biobank subjects that did not carry any rare variant (biobank-specific carrier frequency ≤ 0.1%) in their associated gene(s) (see Methods). We then compared haploinsufficient disease prevalence between these two groups. This analysis was limited to those diseases with at least 1 diagnosis in either the carriers or non-carriers across the two biobanks (*N*=28; 63 diseases either lacked structured diagnostic codes or had no diagnosed cases in either biobank). In a cross-biobank meta-analysis, the pLoF variants had a statistically significant (Bonferroni-corrected) effect on prevalence for approximately two-thirds (20/28) of the diseases (see Supplementary Dataset 4 for the full results). Moreover, the prevalence risk ratios for the significant associations (adjusted for recruitment age, birth sex, and the first 16 principal components of the genetic covariance matrix using logistic regression) were correlated across the two biobanks (*R*^*2*^ = 0.53; *P*-value = 1.27×10^−4^, Figure 2A).

**Figure 2.**
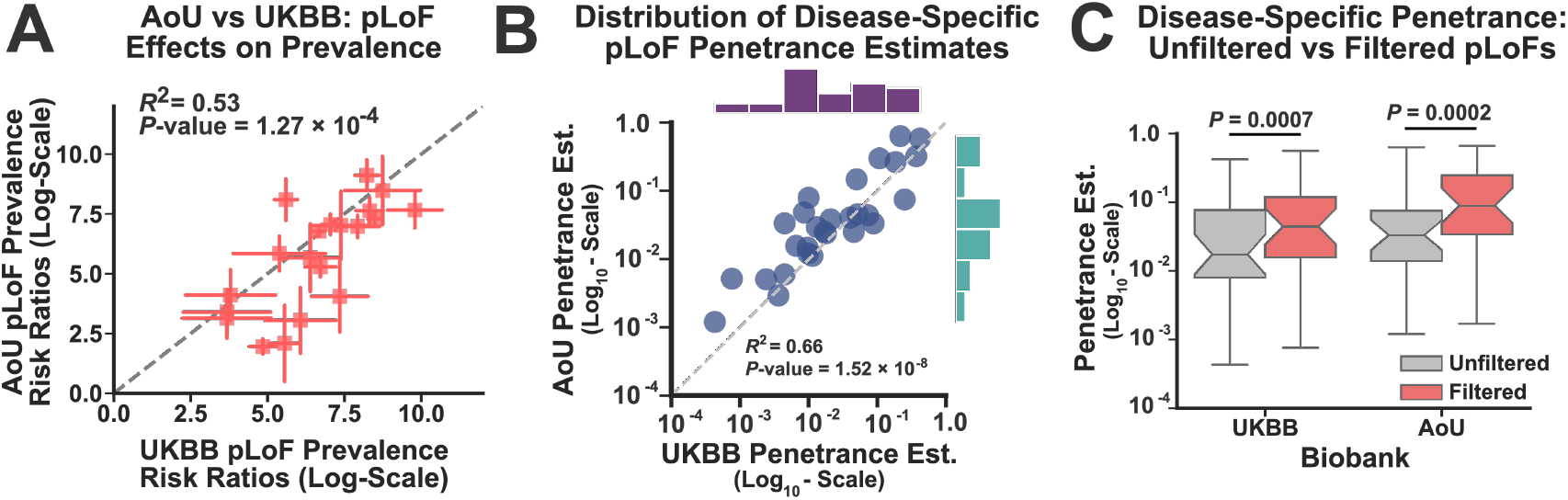
pLoFs Have Detectable Phenotypic Effects but with Reduced Penetrance. (A): Average pLoF effects on disease prevalence are compared across the two biobanks. Note, this panel only displays penetrance risk ratios for those diseases in which pLoFs had a statistically significant effect in the cross-biobank meta-analysis (*N*=20). Datapoints represent the estimated prevalence risk-ratio while bars indicate ± one standard error of this estimate. Correlation was assessed using Pearson’s correlation coefficient (R^2^). (B): Disease-specific penetrance estimates for pLoFs were computed in each biobank using diagnoses only (*N*=28). The estimates from the two biobanks were compared, and their correlation was assessed using Pearson’s correlation coefficient (R^2^). (C): Boxplots comparing the distribution of penetrance estimates before (gray) and after (red) filtering pLoFs for potential artifacts (see main text). Statistical significance was assessed using a one-sided Wilcoxon signed rank test.

These results indicate that pLoFs had consistent effects on prevalence, but this does not imply that their apparent penetrance was high. To estimate pLoF penetrance for each disease, we measured their expression using EHR-derived diagnoses, generating point estimates and 95% confidence intervals for penetrance using a simple binomial model (see Methods). Figure 2B compares the disease-specific pLoF penetrance estimates across the two biobanks (again limited to diseases with diagnostic data available in one of the two biobanks; *N*=28). Clearly, this is an imperfect estimate, as only diagnoses were used to measure disease expression. Nevertheless, the penetrance estimates across biobanks were highly correlated (*R*^*2*^ = 0.66; *P*-value = 1.52×10^−8^), indicating replicability but also highlighting significant disease-wide heterogeneity. Consistent with previous analyses^44^, the pLoF penetrance estimates were relatively low (median across diseases = 1.7% ± median absolute deviation = 1.6% in the UKBB; 3.3% ± 2.2% in AoU).

Most studies that analyze pLoFs filter these variants to remove potential annotation artifacts^52–55^, which are variants that have no molecular impact but were misannotated as LoFs by the variant prediction software. To account for such artifacts, we repeated this analysis after removing variants that impacted non-canonical transcripts (i.e. non-MANE Select^56^) and/or failed to meet a set of quality filters (assessed using the LOFTEE package^53^). In total, this filtering removed 29% and 38% of the pLoFs in the UKBB and AoU respectively (mean ± std. deviation across individual diseases: 35% ± 25% and 42% ± 25%, respectively). Restricting the analysis to the filtered variants increased the disease-specific penetrance estimates in both biobanks (Figure 2C; Wilcoxon Signed Rank Test Meta Analysis *P*-value = 9.54 ×10^−7^); seven diseases even achieved pLoF penetrance estimates exceeding 20% in both datasets (ex: Autosomal Dominant Polycystic Kidney Disease and Neurofibromatosis Type 1; see Supplementary Dataset 5 for details. Note, most individual disease penetrance estimates in AoU must be suppressed due to privacy restrictions). Nevertheless, while the median penetrance estimates across diseases increased nearly 3-fold in both biobanks, they remained less than 10% (4.6% ± 3.7% in UKBB; 9.0% ± 7.2% in AoU).

Therefore, variant annotation artifacts alone unlikely account for a substantial fraction of the reduced pLoF penetrance.

### Reduced Penetrance Persists after Accounting for Missing Disease Diagnoses

Thus far, diagnoses have been used to measure disease expression and estimate penetrance. This is clearly an imperfect approach. Most haploinsufficient (*N*=63) diseases either lack the formal diagnostic data needed to estimate penetrance or simply have no diagnoses available in either biobank. Moreover, even when diagnoses are available, previous work suggests that many pathogenic variant carriers exhibit disease expression without formal diagnoses^46,57^. To overcome this limitation, we also quantified disease expression using covariate-corrected Phenotype Risk Scores (PheRS)^58,59^. These scores measure the extent to which a subject is a phenotypic outlier based on their pattern of expressed, disease-related symptoms. This analysis is possible even in the absence of formal diagnostic data (see Methods).

Figures 3A (UKBB) and 3B (AoU) display the distributions over the median PheRS estimates for each disease, which were standardized using the PheRS distributions observed in non-carriers (Methods). Although the median increase in scores among carriers relative to non-carriers was modest (0.06 in the UKBB; 0.10 in AoU), the PheRS estimates were systematically increased among carriers across the full set of diseases (Wilcoxon Signed-Rank Meta-Analysis *P*-value = 2.88×10^−11^). We also compared the distributions of PheRS’s among the carriers and non-carriers of the individual diseases, using one-sided Brunner-Munzel non-parametric statistical hypothesis tests to assess significance. Multiple diseases achieved dataset-wide (14 of 89 diseases with PheRS information available in both biobanks) or at least uncorrected (37 of 89) statistical significance in a cross-biobank meta-analysis (see Supplementary Dataset 6 for the full set of results). Therefore, the pLoF variants were systematically associated with increased symptom expression for many diseases, at least when compared to a set of biobank-derived non-carriers. However, the increase in scores among the carriers was relatively modest for most except for a few outliers (see Figures 3A and 3B).

**Figure 3.**
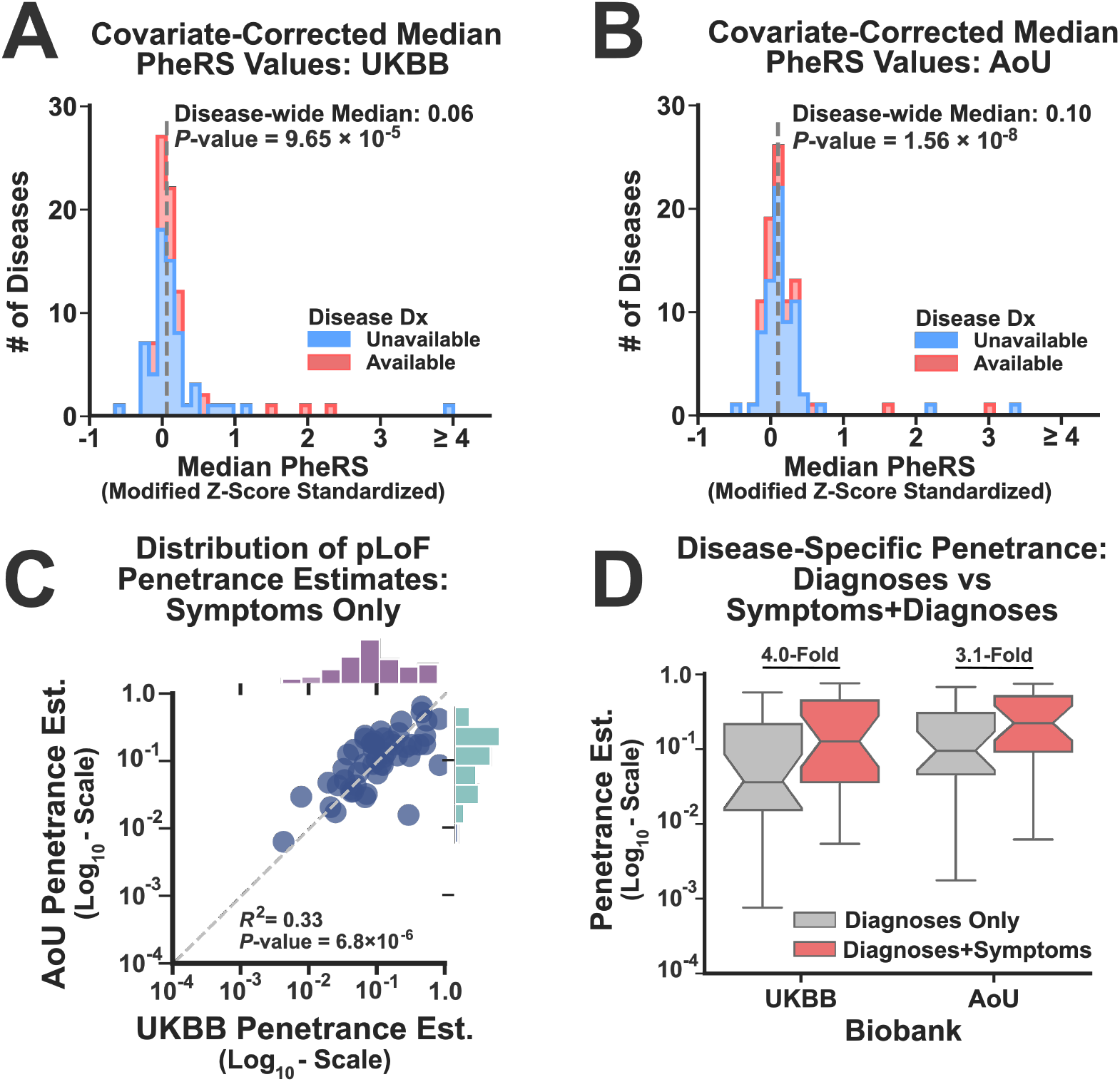
Reduced Penetrance Persists after Accounting for Missing Disease Diagnoses. (A, B): The median Phenotype Risk Score (PheRS) for each disease was computed using the pLoF carriers and standardized using the scores observed in the non-carriers. These histograms display the standardized, disease-specific median PheRS scores for the UKBB (B; N=90 diseases with PheRS information available) and AoU (C; N=90). Significance was assessed using a one-sided Wilcoxon signed rank test. (*C*) For those diseases without diagnostic data (N=51), the symptom-driven, disease-specific penetrance estimates for the pLoF variants were compared across the two biobanks. Correlation was assessed using Pearson’s method (R^2^). (D): For diseases with symptoms and diagnoses available in both biobanks (N=25), the boxplots compare the disease-specific penetrance estimates before (gray) and after (red) incorporating the symptom-driven expression measurements. The fold-increase in the median that occurred after including the symptom-driven measurements is depicted at the top of the boxplots.

Unfortunately, the PheRS method cannot directly be used to produce penetrance estimates. It is a continuous score whose range varies with the number symptoms assigned to each disease. Moreover, the threshold for an elevated PheRS depends upon the background frequency of disease-associated symptoms in the general population of non-carriers. Common symptoms (examples: fatigue, muscle pain, proteinuria, etc.) will yield non-zero PheRS’s but may not be indicative of pLoF expression. To overcome these issues, we developed a statistical model that converted PheRS’s into disease expression probabilities by comparing the frequency of diagnosed symptoms among pLoF carriers to a background distribution. If a carrier’s diagnosed symptoms were unlikely according to this background distribution but were consistent with the symptoms expressed by other pLoF carriers, then they were assigned a high probability of disease expression (see Methods). This model generated a unique expression probability estimate for each pLoF carrier, which could then be incorporated into our penetrance estimates (see Methods). In Figure 3C, we display the distributions of symptom-driven penetrance estimates for the 51 haploinsufficient diseases that lacked diagnostic data but shared documented symptoms across the two biobanks. For these diseases, the purely biobank-specific, symptom-driven penetrance estimates were correlated (*R*^*2*^ = 0.33; *P*-value = 6.8 ×10^−6^), indicating that these measurements were capturing consistent phenotypic effects as well as the penetrance heterogeneity across diseases (see Supplementary Information for additional information).

Overall, incorporating symptom-driven expression measurements increased our sample size and improved our ability to detect haploinsufficient disease expression in the biobanks. However, for diseases with phenotype data that was shared by both datasets (*N*=76), pLoF penetrance remained incomplete for all diseases (median penetrance estimates: 8.4% ± 6.1% and 14.1% ± 8.6% in the UKBB and AoU respectively). For those diseases with both diagnoses and symptom-driven expression measurements available in both biobanks (*N*=25; 3 diseases from Figure 2 did not share symptom data across biobanks), including symptoms increased the penetrance substantially (12.5% ± 11% and 22.0% ± 17.4% penetrance for the UKBB and AoU, respectively, Figure 3D). However, most pLoF carriers still did not display evidence for disease expression. The full set of symptom-driven penetrance estimates (which include diagnosis-derived measurements if available) are provided in Supplementary Dataset 7.

### Incomplete pLoF Penetrance Persists After Accounting for Gaps in EHR Data Coverage

Another factor that could negatively impact penetrance estimation is the incomplete lifetime coverage of the EHR data in biobanks^42,46^. More specifically, these datasets capture only a fraction of their subjects’ lifespans, and the data coverage for individual participants can vary widely (see Supplementary Figure 1). Missing clinical data, which can occur during any lifetime interval (e.g. childhood, late adulthood, etc.), can allow a subject to appear asymptomatic and thus bias penetrance estimates downward. To address this issue, we developed a statistical filtering strategy to remove pLoF carriers whose EHR data coverage was insufficient to accurately measure disease expression (see Methods for details). This filtering removed 17% and 35% of the pLoF carriers from the UKBB and AoU respectively, with more extensive filtering occurring in the latter biobank due to its tendency to have more limited data coverage.

Figure 4A depicts the change in pLoF penetrance estimates (using both diagnoses and symptoms if available, otherwise symptoms only) for the pLoFs within both datasets after removing subjects with low clinical data coverage. Penetrance increased systematically but modestly in both biobanks (fold-increase in median penetrance: 1.9 and 2.0-fold for the UKBB and AoU, respectively; Wilcoxon Signed Rank Test Meta Analysis *P*-value < 2.2×10^−16^). After all three of our data processing steps (annotation artifact removal, accounting for missed diagnoses, and filtering subjects with incomplete clinical data, see Figure 4B), the median penetrance for diseases with diagnoses and symptoms available in both biobanks (*N*=25) increased 10.9-fold (1.7% ± 1.6% vs 18.5% ± 16.7%) and 6.1-fold (3.8% ± 3.2% vs 23.3% ± 19.1%) in the UKBB and AoU respectively (see Supplementary Dataset 8 for the full set of estimates). However, no haploinsufficient disease achieved a pLoF penetrance estimate exceeding 80% in either dataset. Moreover, the median estimates for childhood-onset disorders remained much lower than expected (9.7% ± 6.2% and 16.7% ± 8.9% in the UKBB and AoU, respectively), particularly when compared to the pLoF penetrance for young adult-onset diseases (26.9% ± 22.3% and 41.5% ± 20.0% respectively). Collectively, this suggests that factors other than annotation artifacts and missing data are driving the incomplete pLoF penetrance observed in biobanks.

**Figure 4:**
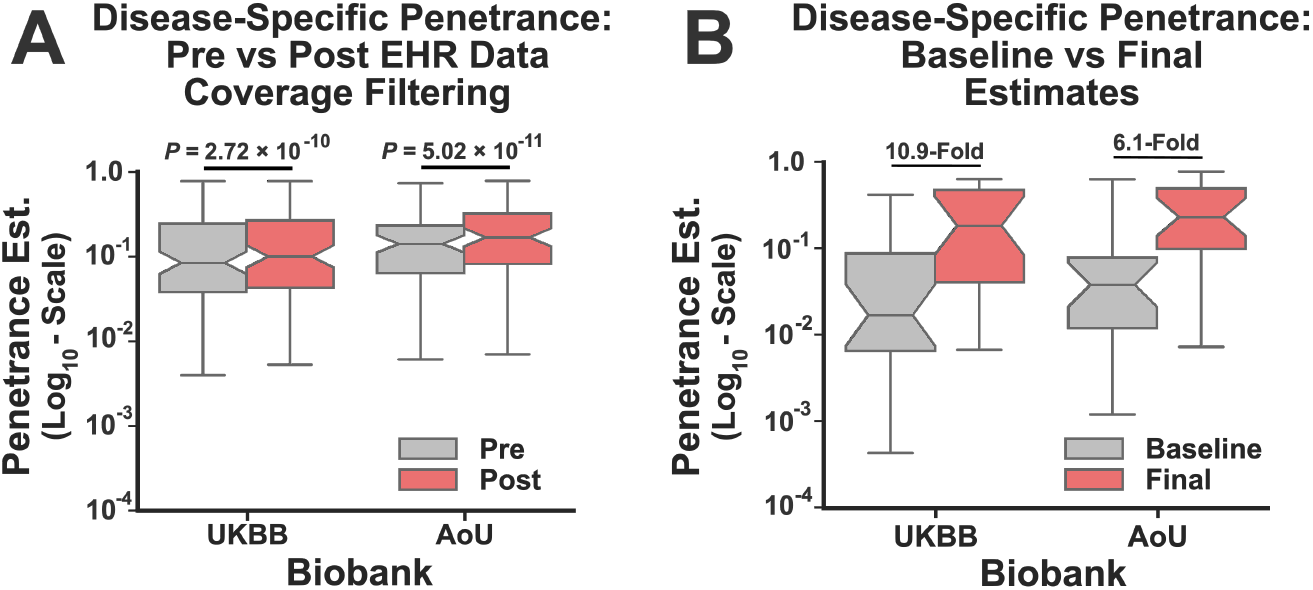
Incomplete pLoF Penetrance Persists After Accounting for Gaps in EHR Data Coverage. (A) These boxplots depict the disease-specific penetrance estimates before (gray) and after (red) removing pLoF carriers with insufficient clinical data coverage. Statistical significance was assessed using a one-sided Wilcoxon signed rank test (*N*=76). (B): For diseases with symptoms and diagnoses available in both biobanks (*N=*25), the boxplots compare the disease-specific penetrance estimates before (gray) and after (red) completing all three of our processing steps. The fold-increase in the median penetrance estimates is shown at the top.

### Variant-Specific Genomic Features Predict Widespread Reduced pLoF Penetrance

After removing annotation artifacts, accounting for missed diagnoses, and correcting for incomplete EHR coverage, pLoF penetrance estimates increased systematically. However, reduced penetrance remained common. None of the filters we employed were perfect, and deficiencies in our processing steps could certainly account for some additional fraction of the incomplete penetrance. However, we also considered the possibility that some pLoFs may simply have intrinsically reduced penetrance. More specifically, it is commonly assumed that variants predicted to cause loss-of-function fully disrupt allelic activity, implying that they are equivalent in terms of their molecular impact (i.e. “complete” loss of function). This may not be the case. Some may have deleterious effects but still harbor residual or even near-complete allelic activity. For example, incomplete non-sense mediated decay escape^60,61^ could allow for the partial expression of a transcript impacted by a stop-gain variant. Such “leaky” expression could in turn cause the apparently reduced penetrance that we observed in the biobanks. This is particularly true for diseases driven by dosage sensitivity, where variability in gene expression and/or activity levels may lead to heterogenous phenotypic expression^62^. Detecting such effects for individual pLoFs using biobank data is challenging. However, by comparing pLoF expression rates to genomic features that may correlate with “leaky” allelic activity, we hypothesized that we could generate systematic evidence for variant-specific incomplete penetrance.

To do so, we conducted a cross-biobank machine learning analysis (illustrated in Figure 5A) designed to predict pLoF penetrance from genomic features that either directly or indirectly capture incomplete loss-of-function. First, we identified cohorts of pLoF variant carriers in both biobanks, filtering out those with inadequate clinical data coverage (see above and Methods). We then measured disease expression among pLoF carriers using symptoms (and diagnoses, if available), producing scores that quantified their expression risk (ranging from 0 to 1, see Methods). Each pLoF was assigned to one of three classes (stop gain, frameshift, or splice change), after which it was annotated with a set of genomic features (6 unique features for stop gain variants, 6 for frameshifts, and 13 for splice changes) predicted to correlate with incomplete loss-of-function (examples are provided in Figure 5A, see Supplementary Information for more details). Finally, using the UKBB exclusively, we trained machine learning models^63^ (one for each pLoF class; see Methods) to predict the pLoF disease expression scores directly from these features. The predictive performance of these models was then independently evaluated in AoU. If our hypothesis were correct, the models should be predictive of pLoF penetrance in this validation dataset, even for variants with prior evidence for pathogenicity according to diagnostic testing laboratories.

**Figure 5:**
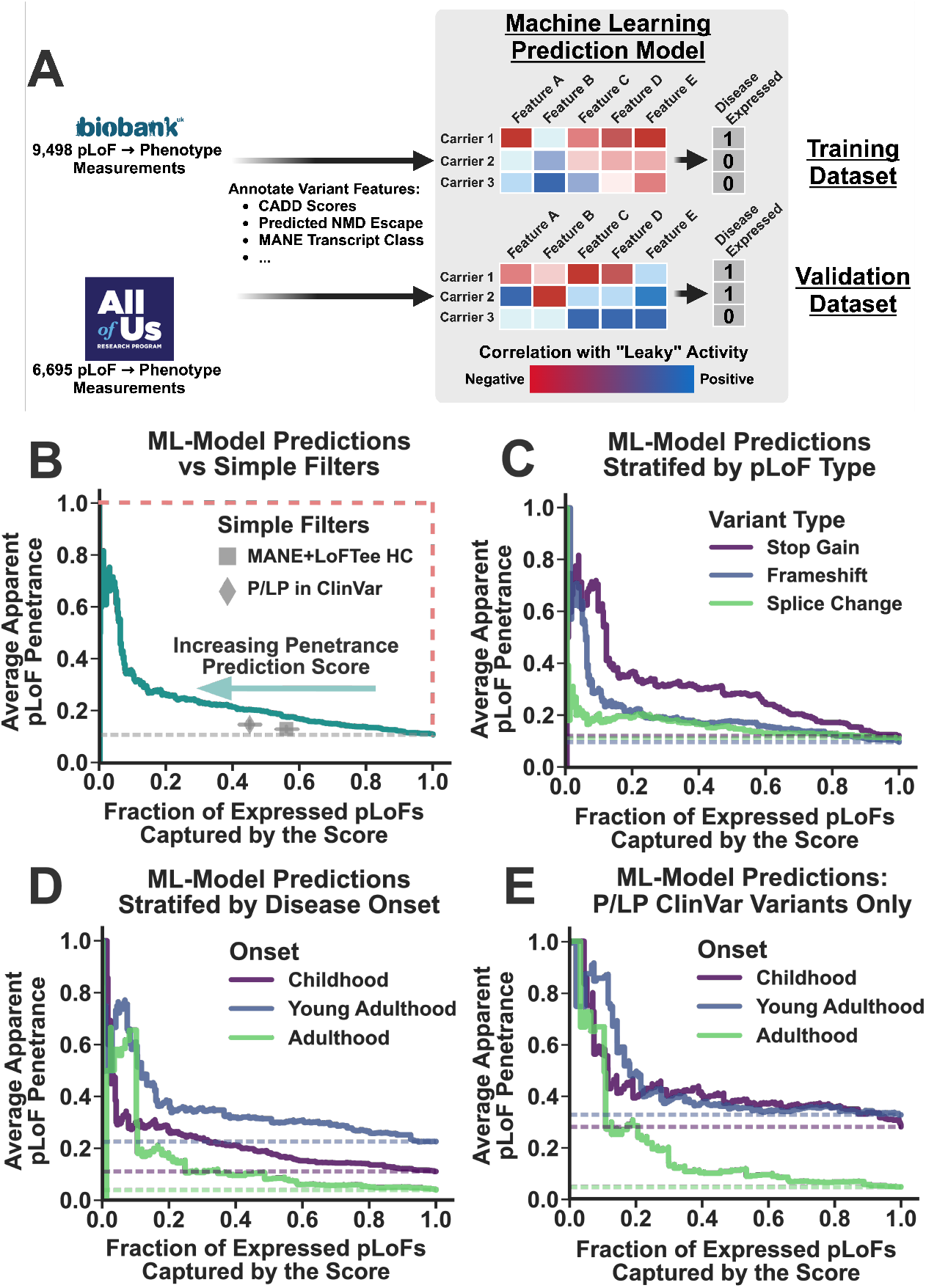
Variant-Specific Genomic Features Predict Widespread Reduced pLoF Penetrance. (A): Schematic illustrating our cross-biobank machine learning analysis to investigate reduced pLoF penetrance. (B): This panel displays the tradeoff between average apparent pLoF penetrance (y-axis) and the fraction of expressed pLoFs recalled (x-axis) when using different model-derived penetrance prediction scores to filter variants. The performance of a simple filter (MANE Select+High-Confidence LOFTEE flag) is shown as a gray square, while a filter that only includes variants with non-conflicting pathogenic/likely-pathogenic (P/LP) annotations in ClinVar is shown as a gray diamond. All error bars represent bootstrapped 95% confidence intervals. The gray dotted line indicates the baseline penetrance rate, and the red dotted line indicates the performance of a perfect classifier (C, D): These panels depict the same results from (B), except now pLoFs are stratified by variant type (C) or typical disease onset (D). Dotted lines indicate the baseline penetrance for each variant or onset class. (E): This panel depicts the penetrance prediction model performance curves for those pLoFs with non-conflicting P/LP annotations in ClinVar, again stratified by disease onset. Created using Biorender.com.

Figure 5B summarizes the performance of the pLoF penetrance prediction models in the AoU validation dataset. Each point on this curve represents a distinct, model-derived penetrance prediction scoring threshold, sorted from highest to lowest. The y-axis depicts the average penetrance for the pLoFs with prediction scores exceeding this threshold, while the x-axis displays the fraction of all penetrant pLoFs (weighted by their disease expression scores) captured by these thresholds (equivalent to recall). Perfect predictive performance is illustrated by the dashed red line (100% average penetrance with 100% recall), while the gray dashed line indicates the baseline penetrance of the pLoFs. If the prediction models are effective, then the average penetrance of the variants should increase systematically as the scoring threshold becomes more selective. Based on the results in Figure 5B, the machine learning penetrance prediction models clearly performed better than baseline (increase in pLoF penetrance from baseline averaged over all possible scoring thresholds^64^ = 11.9%; bootstrapped *P*-value < 1.0×10^−4^), with penetrance exceeding 50% at selective thresholds. For reference, we also depict the performance of two simpler methods for identifying highly penetrant pLoFs: the annotation artifact filter employed previously (pLoFs in MANE Select transcripts^56^ with a high confidence LOFTEE flag^53^) and variants with non-conflicting pathogenic/likely-pathogenic (P/LP) annotations in ClinVar^47^. Averaged over all possible scoring thresholds (while weighting by recall, see Methods), the machine learning models identified pLoFs with significantly higher average penetrance than either (22.7% vs 12.1% or 12.6% respectively; bootstrapped *P*-value < 1.0×10^−4^). Moreover, the models even outperformed these simple filters at the precise thresholds that matched their recall (17.7% vs 13.0% and 20.7% vs 14.8% for MANE Select/LOFTEE and P/LP respectively, see Figure 5B).

In Figure 5C, the performance of the penetrance prediction models is stratified by pLoF class (stop gain, frameshift, and splice change) and compared to the baseline penetrance of each (dotted lines). The models outperformed the baseline penetrance estimates for all three classes (bootstrapped *P*-values < 1.0×10^−4^), but the penetrance for splice change variants was only marginally increased by the model, even at selective scoring thresholds (avg. penetrance increase from baseline: 5.1%). This suggests that this class of variants may be associated with the highest rates of incomplete penetrance. Despite the variability in predictive performance, feature importance analysis^65^ indicated that a consistent set of variant-intrinsic genomic features contributed to the performance each model, with an omnibus score of deleteriousness (CADD^66^) and relative amino acid position^67^ being among the most predictive features for all three (see Supplementary Figures 5-7). That said, there were also some class-specific features that also contributed substantially to predictive performance (ex: features predicting NMD escape for stop-gains^60^, see Supplementary Figure 5), suggesting that penetrance prediction could be improved by incorporating additional class-specific features.

Figure 5D again displays the performance of the penetrance prediction models, except now the results are separated by disease-onset. Even pLoFs in adult-onset, haploinsufficient disease genes can be stratified to maximize penetrance (avg. penetrance increase from baseline = 10.6%; bootstrapped *P*-value < 1.0×10^−4^).

However, they have the lowest average penetrance overall, likely because reduced penetrance is more common for these diseases coupled with the fact their expressivity is the hardest to measure in biobanks due to right censoring. Figure 5E displays the penetrance prediction model performance curves exclusively for variants annotated as P/LP in ClinVar (non-conflicting), again stratified by disease onset. Overall, the machine learning models remained strongly predictive of penetrance for this class of variants (avg. penetrance increase from baseline = 20.0%; bootstrapped *P*-value < 1.0×10^−4^), with the most stringently filtered pLoFs now approaching near complete penetrance for all three onset classes. Based on these results, it is highly unlikely that incomplete clinical data is accounting for much of the apparent reduction in penetrance, as missing data should not correlate with variant-specific genomic features independently of their effects on disease expression. Instead, there is a much more likely explanation for this result^42^: many clinically-annotated P/LP variants have reduced (or even near-zero) penetrance, which can be predicted from genomic features.

## Discussion

Exome and genome sequencing have revolutionized the field of clinical genetics^68^. More rare disease patients are being diagnosed using these technologies, and this has improved our ability to provide timely counseling and treatment to the affected individuals and their families. As a result, there is growing interest in applying broad genomic testing to asymptomatic individuals^7–18^, with the goal of identifying individuals at risk for a Mendelian disease prior to symptom onset. Theoretically, this could lead to better clinical outcomes through earlier diagnosis, surveillance and management. For prognostic genomic testing to succeed, it should at least have a quantifiable positive predictive value^19–21^, a statistic that directly depends on penetrance. Currently, penetrance is almost universally unknown except for a handful of unusually frequent, deleterious variants. As a result, Mendelian disease risk assessments will be imprecise for most asymptomatic carriers. This may have a limited impact on patient outcomes in many settings. However, the clinical decisions made using genomic testing will be life altering in some cases, and without penetrance information, such interventions may be unnecessarily applied to low-risk carriers.

In this study, we investigated the penetrance of one of the simplest classes of clinically relevant genetic variants: predicted loss-of-function variants (pLoFs) in haploinsufficient (autosomal dominant) disease genes. Consistent with prior analyses^43,44,57^, we found that the apparent penetrance of these variants in biobanks was generally low. Accounting for variant annotation artifacts and missing clinical data increased penetrance estimates, but most pLoFs remained unexpressed. Distinguishing between high and low risk pathogenic variants represents a significant challenge for prognostic genetic testing. In diagnostic applications, detailed criteria for variant interpretation have been developed to mitigate the risk for false positive results^28,69^.

These criteria cannot be directly applied to asymptomatic carriers to predict risk. Moreover, even variants with prior evidence for pathogenicity based on diagnoses in symptomatic individuals had incomplete penetrance in the biobanks, suggesting that the utility of clinical annotations for penetrance prediction is limited. Alternatively, machine learning models that incorporated variant-specific genomic features like omnibus scores of deleteriousness^66^, splicing predictions^70^, and putative non-sense mediated decay escape^60^ successfully stratified pLoFs according to their penetrance. This suggests that many pLoFs have (sometimes very) low penetrance, enabling their carriers to avoid disease expression via “leaky” or partial allelic activity.

These results imply that screening tests for and incidental findings related to haploinsufficient diseases that rely on current pLoF interpretation guidelines will have low positive predictive values. However, there are several limitations to this analysis. First, biobanks recruit subjects from adult populations, which are almost certainly depleted of severe, early-onset phenotypes. As a result, pLoF penetrance estimates derived from biobanks may be lower than those obtained using cohorts recruited at birth, although this will vary based on a disease’s age-of-onset, it’s impact on survival, and the recruitment strategies used by the biobanks. Second, our machine learning penetrance prediction models did not incorporate any gene or disease-level features. Penetrance likely varies systematically across these entities, and future work will be needed to decompose their contributions to penetrance. Finally, penetrance variability is also driven by variant-extrinsic features, such as polygenic background and environmental modifiers, and these factors could also be incorporated into the risk prediction models for asymptomatic carriers.

Along these same lines, we limited our analysis to pLoF variants linked to 91 haploinsufficient diseases. Although they covered a range of human pathophysiology, these represent the simplest class of pathogenic variants and capture only a tiny fraction of Mendelian disease-causing mutations. That said, loss-of-function is also the predominant mechanism for hundreds if not thousands of autosomal recessive diseases, and our findings likely generalize to these and other diseases that are sensitive to allelic activity and gene “dosage.” There are also many more diseases (autosomal dominant or otherwise) driven by a variety of molecular mechanisms, and it’s possible that reduced penetrance is even more pervasive for some (ex: missense variants in haploinsufficient disease genes). Finally, the machine learning models used in the current study were developed to provide evidence for widespread reduced penetrance in biobank datasets. However, their accuracy in real-world applications remains unknown. To be clinically useful, much work will be needed to build models that are replicable, calibrated, minimally biased, and broadly applicable to diverse genes, diseases, and populations.

For now, it may be possible to predict the phenotypic outcomes for some rare genotypes with near complete penetrance^71^. In addition, when variant information is combined with orthogonal data like enzymatic activity and biomarkers, the prognostic accuracy may be very high^72^. Unfortunately, such assays are available for only a tiny fraction of genetic diseases. For most, the variants themselves provide the only piece of prognostic information. Prior clinical evidence for pathogenicity may increase the positive predictive value for a particular variant, but based on the analyses presented here, prior pathogenic annotation labels are at best poorly correlated with high penetrance. Given the distinct goals of diagnostic and prognostic genetic testing, this is not unexpected. Access to high-quality outcome data for individual genotypes will certainly help, but given their intrinsically low frequency, it will likely remain difficult to estimate penetrance and predict disease outcomes in individual patients for the foreseeable future. Therefore, we suggest that caution be used when returning positive genomic findings to asymptomatic patients, including those detected incidentally during routine diagnostic testing^18^. Even with prior evidence for pathogenicity, variant-associated disease risk remains uncertain and may be quite low.

## Methods

### Haploinsufficient Disease Curation and Annotation

We used the ClinGen^48^ Database (downloaded on July 25th, 2023) to identify Mendelian disorders that have strong evidence to support haploinsufficiency as a mechanism of disease (ClinGen Dosage Haploinsufficiency Assertion Evidence Level 3). We then aligned these diseases to the Online Mendelian Inheritance in Man^49^ database (downloaded on February 23, 2023) using simple string matching followed by manual curation. This yielded 91 autosomal dominant/pseudo-autosomal dominant diseases linked to 117 genes, which were manually annotated with their typical onset (Childhood, Young Adulthood, Adulthood) and general classification (Congenital Malformation, Isolated Neurodevelopmental, Complex Neurodevelopmental, Tumor Predisposition, and Other) by a board-certified medical geneticist (author D. Blair) using clinical expertise and literature review. Afterwards, disease-specific diagnostic codes were annotated to these diseases by manually curating the terminologies^73^ used by the Observational Medical Outcomes Partnership Common Data Model^74^ (OMOP-CDM).

Finally, the diseases were annotated with a set of Human Phenotype Ontology^75^ (HPO) symptoms using the data from several ontologies, including the HPO itself (downloaded on February 21, 2023), the Disease Ontology^76^ (downloaded on February 23, 2023) and OrphaNet^77^ (downloaded on February 23, 2023 using the HOOM^78^ module). The sequence and transcript information for each of the 117 genes was downloaded from the Ensembl^79^ database (Release 109; GRCh38 assembly) using the PyEnsembl^80^ package. Additional gene and transcript information (exon-intron boundaries, 5’ and 3’ UTRs, full coding and amino acid sequences) was downloaded using gget^81^. The 91 haploinsufficient diseases, along with their annotated information, are provided as Supplementary Dataset 1.

### Aligning HPO Symptoms to the OMOP-CDM Terminology

To identify HPO^75^ symptom expression in the EHR data, we needed to align this ontology to the structured diagnostic data available in the electronic health records of each biobank. Because both biobanks encode their clinical data using the OMOP-CDM^74^, we focused on aligning the HPO symptom terminology to the structured vocabulary used by this data model^73^. Unfortunately, aligning the HPO to other medical terminologies is largely an unsolved problem that lacks a consensus regarding best practices^82^. Therefore, we created a custom alignment by building on our previous work^25^ while implementing some new techniques.

First, we created an alignment map between the HPO and SNOMED-CT^83^, as the latter represents the most comprehensive medical terminology available for the dissemination of EHR data. It is also fully incorporated into the concept terminology used by the OMOP-CDM. To create an HPO-to-SNOMED map, we followed the approach of McArthur et al.^84^, who created a similar map between the HPO and PheCodes^85^. First, we constructed a map linking HPO to SNOMED-CT terms if they shared a common concept in the UMLS Metathesaurus^86^. Second, we used an ontology alignment algorithm (SORTA^87^) to find all SNOMED-CT terms that mapped to an HPO term with a similarity score of ≥ 0.8 for at least 1 of their associated string pairs (both SNOMED-CT and HPO often provide multiple strings for each term). For terms with multiple aligned string pairs, we collated all the similarity scores across the different string pairs, storing both an average and maximum score.

With an HPO-to-SNOMED map in place, the HPO terms themselves could be directly aligned to the concept terminology used by the OMOP-CDM, as a map from SNOMED-CT terms to the OMOP-CDM concepts is provided by the Observational Health Data Sciences and Informatics (OHDSI) Collaborative^73,88^. However, it is important to note that one HPO term often mapped to multiple SNOMED-CT terms, which could then map to the OMOP vocabulary in multiple ways. Therefore, each HPO-OMOP alignment was often supported by multiple intermediary relationships. To summarize this phenomenon, we stored several pieces of information for each alignment that captured the quality of its supporting evidence. These included: the total number of intermediate relationships supporting the mapping, the fraction of these relationships that were supported by the UMLS, the fraction that achieved a SORTA string alignment similarity score ≥ 0.8, the average SORTA score across intermediaries, and the maximum score achieved. In total, this process generated 35,825 unique HPO-to-OMOP alignments.

Because automated alignments like this tend to be rife with spurious results, one of the authors (D. Blair) manually reviewed 500 random mappings and annotated their medical accuracy. The accuracy was unsurprisingly variable, but overall, far better than random (average precision: 0.76). To further improve accuracy, we built a simple logistic regression classifier (implemented in sklearn^89^) to predict if an HPO-OMOP alignment was accurate. The model incorporated the alignment features described above as linear predictors (noting that the maximum achieved SORTA score was incorporated as interaction term with the total number of intermediate relationships). The model was trained on the 500 manually annotated alignments prior to being applied to the full dataset. In leave-one-out 5-fold cross validation experiments, the area under the receiver operator characteristic curve for the model predictions was 0.76 (standard error: 0.017), indicating that these predictions could provide a substantial improvement to alignment accuracy. Therefore, all ≈35,000 HPO-to-OMOP alignments were scored using the prediction model, and several false positive rate (FPR) thresholds were selected for downstream filtering. The complete set of HPO-to-OMOP Concept ID alignments (along with their features, manual annotations, machine learning scores, and whether they survived various FPR filtering thresholds) are provided as Supplementary Dataset 9. Finally, we experimented with various alignment FPR thresholds in downstream analyses. Overall, PheRS enrichment among pLoF carriers was highest when using the relationships that survived a 20% FPR threshold (data not shown).

Therefore, this set of alignments was used for the results reported in thes manuscript.

### Sequence Data Quality Control, Variant Annotation, and Non-Carrier Cohort Identification

This study utilized the exome sequence (ES) data from the UK Biobank (UKBB)^37^ and the genome sequence (GS) data from the All of Us (AoU) Research Program^41^ to investigate the penetrance of predicted loss-of-function (pLoF) variants in haploinsufficient disease genes. For the AoU dataset, the GS samples undergo an extensive quality control process, which ensures that samples meet several coverage and accuracy thresholds^41^. Therefore, all samples with GS data that were not flagged by AoU’s quality control pipeline were analyzed in this study (*N* = 245,376). For the UKBB, less sample-level quality control was performed *a priori*. Therefore, the ES data from this biobank underwent additional quality control filters consistent with those performed in previous studies^90^. Briefly, all samples that showed evidence for genetic and self-reported sex discordance (*N* = 296), sample duplication (*N* = 56), excessive SNP array-short read sequencing genotype discordance (*N* = 513, including those that lacked array data), low read coverage for the haploinsufficient genes of interest (20x coverage at less than 90% of the base pairs; *N* = 92), and excessive missing genotypes (*N* = 329, again limited to the haploinsufficient genes of interest) were excluded from the analysis (total number of samples that failed quality control: 1,156). After excluding subjects that withdrew from the UKBB study, this dataset contained a total of 468,672 subjects with both ES and EHR data.

Following sample-level quality control filtering, variants from the exonic regions of the haploinsufficient disease genes were isolated from both datasets (performed using bcftools^91^ in the UKBB and hail^92^ in AoU), storing the variant genotyping calls in VCF files. Individual-level data was then stripped from these files, and the predicted molecular effect of each variant was annotated using VEP^93^ (Version 110).

Simultaneously, the variants were annotated with any previous interpretations documented within the ClinVar^47^ database (downloaded on May 13^th^, 2024). Finally, all pLoFs within these datasets were identified using the LOFTEE^53^ plug-in for VEP, which also provided a flag indicating the overall confidence in this assessment (high vs low confidence; HC vs LC). Using the output from VEP, each pLoF was annotated with its most clinically significant impacted transcript^56^ (MANE Select, MANE Plus Clinical, Other), and the variants were also assigned to one of three pLoF classes: stop gain, frameshift, and splice change, with other variant types (e.g. missense) excluded.

Following annotation, we returned to the VCF files that contained the individual-level genotype calls and isolated all pLoFs identified in the previous step. We then identified all carriers for each individual variant, removing those that did not meet a basic set of genotype-specific quality control filters^90^. For single nucleotide variants (SNVs), we assigned a no-call status to all carriers with a genotyping quality score < 30, sequencing depth < 7, or alternate allelic balance < 0.15. For insertion-deletion variants, we were more stringent and removed those calls with a quality score < 30, sequencing depth < 10, or alternate allelic balance < 0.2. In addition, we removed a variant from the analysis entirely if its call rate was <0.99 or if its carrier frequency in the biobank was greater than 0.1% (to remove common variants that were more likely to have low or no penetrance). For the UKBB, we also *a priori* removed those variants that achieved an average read depth <10 for more than 10% of the samples in the dataset (per recommended best practices^94^). In total, this process identified 3,131 (Supplementary Dataset 2) and 3,889 (Supplementary Dataset 3) pLoFs carried by 14,010 and 11,022 subjects in the UKBB and AoU respectively. Note, some individuals harbored multiple pLoF variants within a single gene (*N*=637 in the UKBB; *N*=304 in AoU), suggesting the possibility of sequencing artifacts versus *in cis* rescue events. Such carriers were not excluded from basic pLoF carrier frequency estimates (i.e. Figure 1) but were excluded from all other analyses. As a result, the impact of *in cis* rescue events on penetrance is a target for future work^55^. Individuals with multiple pLoFs in distinct halploinsufficient disease genes were not dropped from the analysis.

Finally, for each haploinsufficient disease, we created a unique cohort of non-carriers that were unlikely to be at risk for the disease of interest. To do so, we first identified all subjects in both biobanks that carried any rare variant (allele frequency ≤ 0.1%) in the gene(s) annotated to each disease. We then dropped these subjects from the set of all biobank subjects (along with those that had a no-call genotype at one of the detected pLoFs). This yielded a unique set of non-carriers for each disease. The total number of non-carriers available for the different diseases was variable but exceeded 280,000 and 130,000 in all instances for the UKBB and AoU respectively.

### Haploinsufficient Disease Prevalence and Penetrance Analysis

The simplest way to measure pLoF-driven disease expression was using diagnoses. Let 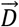 denote a binary vector of length *N*, where in *N* is the number of pLoF carriers for some haploinsufficient disease of interest plus the number of non-carriers identified in the biobank. Let *D*_*i*_= 1 denote that the *i*th subject was diagnosed with the disease of interest at least once in their EHR data, where diagnoses were identified using a set of manually annotated OMOP-CDM concept codes (see Supplementary Dataset 1). Finally, let 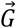 denote a binary vector indicating the pLoF carrier status for the *N* subjects. We estimated a biobank-specific pLoF prevalence risk ratio for each disease (on the logarithmic scale and approximated using the log-odds ratio; denoted *γ*_pLoF_) using one of two approaches. For the more common diseases 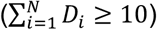, we incorporated covariates into the analysis using the following log-linear model:

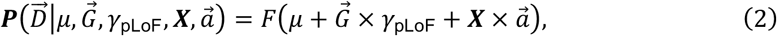

where *F* denotes the logistic function, *μ* is an intercept term, ***X*** is a matrix of covariates, and 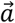 is a vector of covariate effect size parameters. For the current study, we incorporated the following covariates into our analysis: recruitment age, birth sex, and the first 16 principal components of the genetic relatedness matrix. Model fitting was performed using Firth-penalized maximum-likelihood estimation^95^, and statistical inference was conducted using a likelihood ratio test. Even with Firth penalization, model inference returned spurious results when 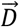 was extremely sparse. Therefore, for very rare diseases 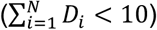, we constructed 2 × 2 contingency tables from 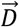 and 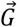 and estimated the (covariate-uncorrected) pLoF prevalence risk ratio and its corresponding standard error using the statsmodels^96^ software package in Python. *P*-values were computed using Fisher’s exact test^97^. Finally, we performed cross-biobank meta-analyses of the pLoF prevalence risk using the Cochran-Mantel-Haenszel Test for stratified contingency tables (implemented in the statsmodels^96^ package).

Supplementary Dataset 4 contains a summary of the results of our disease-specific prevalence association analyses.

To estimate disease-specific pLoF apparent penetrance (denoted 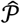) using diagnoses, we assumed a symmetric beta prior distribution over 𝒫 with hyper-parameter *θ* = 0.5. Assuming disease diagnoses among pLoF carriers follow a Bernoulli process, the posterior distribution over 𝒫 is:

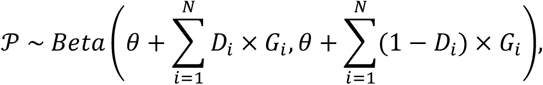

such that an estimator for the mean pLoF penetrance (denoted 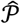) is simply:

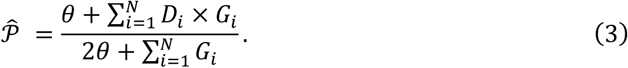

In practice, disease-specific penetrance was estimated in each biobank independently, allowing the estimates to be compared across datasets (ex: Figure 2B). Note, the prior distribution was incorporated into the analysis to smooth our penetrance estimates in the face of sparse data. For diseases with very low pLoF carrier counts, the prior can have an outsized effect. This should have no appreciable impact on our machine learning analysis but may upwardly bias absolute pLoF penetrance estimates, especially when the number of pLoF carriers is small.

### Phenotype Risk Score Analysis

Most haploinsufficient diseases lack structured diagnostic codes that can be used to identify disease expression in EHR data. In such instances, it can be difficult to determine if a subject is in fact expressing the disease phenotype. Phenotype Risk Scores^58,59^ (PheRS’s) were developed to address this issue. These scores measure the extent to which a subject represents an outlier in phenotype space. Their effectiveness relies on a critical assumption: affected Mendelian disease-associated variant carriers should express constellations of symptoms that are highly atypical when compared to their non-carrier counterparts. Although this is sometimes true, it is not the case for all diseases. Moreover, non-carriers can become phenotypic outliers as well, for example, if they develop unusual complications from a common disease or multiple diseases at once. Therefore, PheRS’s represent an alternative, albeit imperfect, method for measuring disease expression among pathogenic variant carriers.

Let 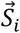 denote a binary vector of length *K*, where is *K* is the number of symptoms annotated to the haploinsufficient disease of interest (derived from multiple ontologies and aligned to structured EHR data, see above). Let *S*_*ik*_ = 1 indicate that the *k*th symptom was diagnosed at least once in the *i*th subject’s EHR data. Finally, let 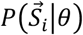 denote a population-wide, background probability model (defined by the parameter set *θ*) that yields the risk of observing the symptom set 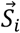. The Phenotype Risk Score for the *i*th subject (denoted PheRS_*i*_) is given by:

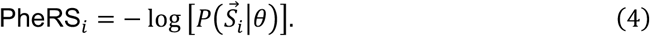

This formula is equivalent to the surprisal, or information content, of the diagnosed symptom set according to the model defined by *θ*, and it provides a measurement for how unusual or atypical this set of diagnosed symptoms is. For the approach to be effective, we must define the background symptom expression probability model.

Consistent with prior studies^58,59^, we assumed a very simple (and somewhat unrealistic) background model for the observed symptoms sets: each of the *k* symptoms occurs independently of the others according to a Bernoulli process. Therefore,

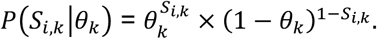

To estimate the background model parameters, we assumed that the Mendelian disease cases were sufficiently rare in the general population such that their risk of biasing the symptom-specific parameter estimates (denoted 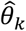) was negligibly low. Therefore, we estimated each symptom expression parameter independently using the maximum likelihood estimator for a Bernoulli process:

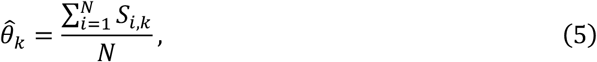

where *N* denotes the total number of subjects in the biobank. After estimating this expression model, the PheRS_*i*_ for each subject becomes:

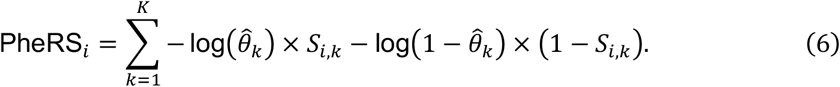

In practice, we further adjusted the raw PheRS’s for confounding covariates (recruitment age, birth sex, and the first 16 components of the genetic relatedness matrix) using ordinary least squares regression.

After computing the covariate-adjusted, disease-specific PheRS’s for every subject in both biobanks, we compared the distributions of these scores among pLoF carriers to those of their non-carrier counterparts. To asses statistical significance, we used a one-sided Brunner-Munzel Non-Parametric Hypothesis Test (implemented in scipy^97^), which evaluated the alternative hypothesis that the PheRS’s observed in the pLoF carriers were stochastically greater than those observed in non-carriers. A fixed effects meta-analysis was used to assess cross-biobank significance. Finally, for the histograms in Figures 2B and 2C, the median PheRS’s in the pLoF carriers were converted to modified Z-scores using the medians and median absolute deviations estimated among the non-carries. The complete set of PheRS results for all diseases are provided in Supplementary Dataset 6.

### Measuring Symptom-Driven Disease Expression

PheRS’s can suggest that a subject is a phenotypic outlier, but these scores do not necessarily provide an accurate measurement of whether a Mendelian disease is being expressed or not. For example, consider autosomal dominant polycystic kidney disease (ADPKD). Clearly, a pLoF carrier who has bilateral renal cysts complicated by chronic kidney disease is expressing the phenotype, but what if a carrier only experiences proteinuria? Proteinuria is certainly a symptom of ADPKD, so this carrier’s PheRS score will be greater than 0. But proteinuria is an extremely common symptom in the general population. Therefore, just because an ADPKD pLoF carrier experiences proteinuria at some point in their life doesn’t mean that they are expressing ADPKD.

To overcome this issue, we formulated the following probability model to measure symptom-driven disease expression among the pLoF carriers. Rather than relying on disease diagnoses, the model uses the pattern of symptoms documented within the EHR data to estimate disease expression risk. Disease diagnoses were specifically excluded from the set of possible symptoms. Therefore, this approach represents an orthogonal method for measuring haploinsufficient disease expression using EHR data. Consistent with the PheRS approach, let 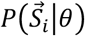 denote the probability that a set of symptoms 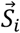 is being expressed according to some general background distribution. In addition, let 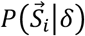 denote the probability that this symptom set is instead expressed by an affected haploinsufficient disease variant carrier (where the parameter set *δ* defines the expression model). Finally, let *E*_*i*_ = 1 indicate that the disease of interest was expressed in the *i*th carrier. The probability of disease expression is given by:

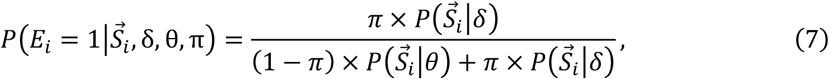

where *π* is the prior probability of disease expression among all carriers. An illustration of this model is provided in Supplementary Figure 2.

Ultimately, our goal in specifying this model was to estimate 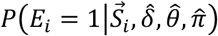, which is the probability of disease expression in the *i*th pLoF carrier conditional on their observed symptoms (denoted 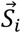) and the inferred expression model parameters (denoted 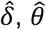 and 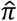). Our procedure for model inference and subsequent expression probability estimation is described in detail in the Supplementary Information. Following estimation, we directly incorporated these symptom-driven expression measurements into our estimates of disease-specific apparent penetrance using the following formula:

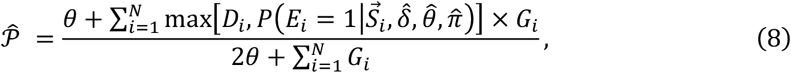

where max[*A, B*] simply returns the maximum of *A* and *B*. Note, when a disease lacked EHR-derived diagnostic data, *D*_*i*_ was simply set to 0 for the relevant pLoF carriers.

### Strategy for Removing pLoF Carriers with Incomplete Clinical Data Coverage

The EHR can provide an incomplete representation of a patient’s disease expression. Subjects are enrolled into biobank cohorts well into adulthood, and there is no guarantee that the records captured by the study represent their complete clinical history. Moreover, the clinical trajectories for many subjects are right-censored, meaning that they have yet to reach the target endpoint for the analysis of interest (in this case, death or disease expression). These gaps in clinical data coverage could account for much of the incomplete penetrance that we observed. Therefore, we devised a method to flag and remove subjects from our analysis whose clinical data coverage was insufficient for reliable measurements of disease expression.

Let 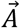 denote a binary vector of asymptomatic indicators, where *A*_*i*_ = 1 indicates that the *i*th pLoF carrier had no evidence for disease expression based on disease-specific diagnostic code(s) and/or documented symptoms (i.e. had no disease-relevant diagnoses or symptoms). Moreover, let ***W*** denote a matrix of clinical data coverage statistics. For this analysis, we used the following four statistics to define data coverage: Age at First Clinical Encounter, Age at Recruitment, Total Number of Documented Clinical Encounters, and Age at Last Clinical Encounter (see Supplementary Figure 1 for their distributions). These four statistics provided a basic summary of a patient’s interaction with the healthcare system, at least according to the information in the biobanks. Finally, let 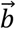 denote a vector of coverage statistic effect size parameters. We modeled the probability of phenotype *non*-expression (i.e. asymptomatic status) conditional on clinical data coverage using the following log-linear model:

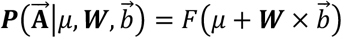

where *F* denotes the logistic function and *μ* is the intercept term. Because different diseases will have different coverage requirements (depending on their typical onset, length of observation time needed to document symptoms, etc), we fit three versions of this model in both biobanks by grouping diseases based on their typical onset (childhood, young adulthood, and adulthood, see Supplementary Dataset 1).

More specifically, each model was repeatedly fit in both biobanks using leave-one-out 5-fold cross validation (model fitting was performed using the LogisticRegression function available in sklearn^89^ with default hyperparameters). For each iteration, 80% of the onset-grouped pLoF carriers were used to estimate the parameters for the disease non-expression model. The remaining 20% were used for validation. Model performance was assessed in the validation subset using the area under the receiver operating characteristic curve (see Supplementary Figure 3 for results). To flag pLoF carriers with insufficient clinical data, we identified the 5% false positive rate threshold in each validation subset. If a subject in a validation subset had a non-expression probability that exceeded this threshold, they were deemed to have insufficient data for disease expression measurement and were flagged for removal in downstream analyses.

### Predicting pLoF Penetrance using Variant-Specific Features

We hypothesized that some pLoF variants have intrinsically low penetrance, which is driven by residual allelic activity. If true, then it should be possible to stratify pLoFs according to their penetrance risk using genomic features that correlate with incomplete loss-of-function. To test this, we developed machine learning models that predict pLoF penetrance risk given a set of genomic features that may correlate with residual allelic activity. Let 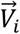 denote the vector of genomic features that characterize the penetrance risk for the pLoF carried by the *i*th subject. Examples of such features include its relative position within the amino acid sequence^67^, the probability that it disrupts splicing^70^, or an omnibus score summarizing its “deleteriousness”^66^. The goal of this analysis is to predict the disease expression risk for the *i*th pLoF carrier using these features:

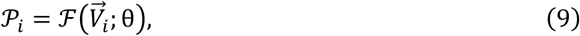

where 𝒫_*i*_is the disease expression score in the *i*th subject (i.e. 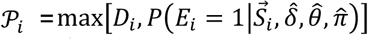, see Eqn. 8) and ℱ is some function that maps the vector 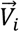 onto disease expression probability space (via a parameter set *θ*). Practically, several different models can accomplish this goal. For this study, we constructed ℱ using the random forest algorithm^63^ implemented in sklearn^89^, which builds predictive models via an ensemble of individual decision trees. Note, we also considered simpler methods for constructing ℱ (i.e. penalized logistic regression) but found that they performed systematically worse than this ensemble learning strategy (see Supplementary Figure 4), likely due to the latter’s ability to capture non-linear effects.

Different types of pLoF variants have distinct features that likely impact their residual allelic activity and thus risk for expression^54^. Therefore, distinct penetrance prediction models were constructed for each of the three variant types analyzed in this study (stop gains, frameshifts, and splice changes). The features incorporated into each of our random forest prediction models are described in detail in the Supplementary Information. Model inference was performed by minimizing the logarithmic loss function of the prediction model when applied to a training cohort of carriers (training algorithm hyperparameters: min_samples_leaf=5, min_samples_split=10, n_estimators=500).

Training was performed exclusively within the UKBB using a subset of pLoF carriers whose disease expression measurements were binarized to simplify inference (see Supplementary Information for details).

After fitting the machine learning models in the UKBB, they were independently validated within AoU. Model performance was assessed by ranking pLoFs according to their penetrance prediction scores (largest-to-smallest). Afterward, we computed the average penetrance (denoted as 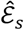 for the *s*th scoring threshold) and the fraction expressed pLoFs (weighted by their expression scores) captured each threshold (equivalent to recall, denoted ℛ_+_) according to the following two formulas:

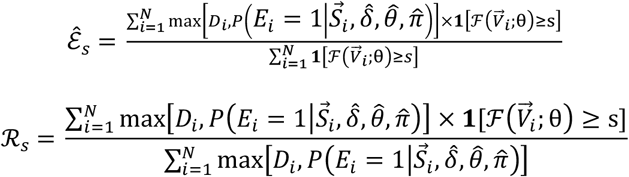

where 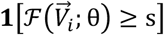 is an indicator function that returns 1 if the score of the *i*th variant is greater than the *s*th threshold. In the machine learning literature, this is equivalent to constructing a precision-recall curve. To measure global model performance, we computed the average penetrance score, which integrates the average penetrance values over all scoring thresholds while weighting them by the change in recall from the previous threshold:

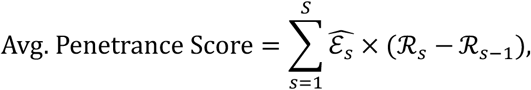

where {*s* = 1 … *S*} denotes the *S* unique prediction scoring thresholds (sorted from largest-to-smallest). This measurement is equivalent to the average precision score, a statistic used by the machine learning community to summarize the performance of prediction models based on their precision-recall curves^64^. Statistical inference for the average penetrance scores (i.e. hypothesis testing and 95% confidence intervals) was completed using bootstrapped resampling^98^. To compare machine learning model performance to the simple filters (MANE Select+High-Confidence LOFTEE Flag or P/LP in ClinVar), the average penetrance score for each filter was computed by treating them as prediction models with only two output values (1: variant passes filter; 0: variant does not pass filter). Note, prediction model feature importance was evaluated within the AoU validation dataset (using Shapley statistics^65^), the results of which are summarized in Supplementary Figures 5-7.

Given that the UKBB and AoU were recruited from the populations of two different countries, the risk that the two datasets contained overlapping individuals (i.e. validation dataset contamination) was very low. However, the two biobanks did contain redundant (variants that occurred multiple times in each biobank) and shared (variants that occurred in both biobanks; *N*=773) pLoFs. Neither were excluded from the training or validation analyses given that they were carried by distinct individuals with conditionally independent phenotypic outcomes. Although this could limit model generalizability, it does not detract from the primary aim of the analysis, which was to demonstrate that pLoFs have intrinsically reduced penetrance. Moreover, Supplementary Table 1 provides a direct comparison of machine learning model performance for all variants and those that were unique to the AoU dataset. The difference in model performance on these two variant sets was minimal (average penetrance score = 22.7% for all variants versus 21.3% for variants unique to AoU).

### Additional Statistical Methods

Unless otherwise noted, the statistical analyses described in the main text and/or figure legends were performed using the implementations (sometimes with slight modification) available in the following Python packages: scipy^97^, sklearn^89^, statsmodels^96^, and pandas^99^. Bootstrapped hypothesis testing was performed by generating empiric distributions for the target parameter estimates using re-sampling with replacement (10,000 re-samples for all tests). Meta-analyses for the different tests were performed using a fixed-effects model based on a standard normal approximation to their test statistics^100^.

## Data Availability

The genomic and electronic health data used for this analysis are publicly available but have strict data use agreements. The process for obtaining access to these biobanks can be found on their respective websites: https://www.researchallofus.org/register/ (All of Us Research Program) and https://www.ukbiobank.ac.uk/enable-your-research/register (UK Biobank). Haploinsufficient disease annotations are provided in Supplementary Dataset 1. The custom HPO-to-OMOP concept alignments generated in this study are provided as Supplementary Dataset 9. All other databases used in this analysis are freely available in the public domain.

## Software Availability

All software packages used to conduct this study are open source and available in the public domain. Phenotype Risk Scores (PheRSs) and symptom-driven disease expression estimates were generated using the SymptomSetModel Python package, which was specifically constructed for this study (https://github.com/daverblair/SymptomSetModel). The scripts used for the analyses described in the paper are also available via github^101^.

## Acknowledgements

The All of Us Research Program is supported by the National Institutes of Health, Office of the Director: Regional Medical Centers: 1 OT2 OD026549; 1 OT2 OD026554; 1 OT2 OD026557; 1 OT2 OD026556; 1 OT2 OD026550; 1 OT2 OD 026552; 1 OT2 OD026553; 1 OT2 OD026548; 1 OT2 OD026551; 1 OT2 OD026555; IAA #: AOD 16037; Federally Qualified Health Centers: HHSN 263201600085U; Data and Research Center: 5 U2C OD023196; Biobank: 1 U24 OD023121; The Participant Center: U24 OD023176; Participant Technology Systems Center: 1 U24 OD023163; Communications and Engagement: 3 OT2 OD023205; 3 OT2 OD023206; and Community Partners: 1 OT2 OD025277; 3 OT2 OD025315; 1 OT2 OD025337; 1 OT2 OD025276. In addition, the All of Us Research Program would not be possible without the partnership of its participants.

This research was conducted using the UK Biobank Resource under Application Number 99922, which relies on data provided by patients and collected by the NHS as part of their care and support. We are extremely grateful to the participants of the UK Biobank, without whom this research would not have been possible.

This work was supported by grants from the National, Heart, Lung and Blood Institute (K38HL164956) and the George Banks and Sarah Ellen Huntington Memorial Fund.

## References

1. Srivastava S, Love-Nichols JA, Dies KA, Ledbetter DH, Martin CL, Chung WK, Firth HV, Frazier T, Hansen RL, Prock L, Brunner H, Hoang N, Scherer SW, Sahin M, Miller DT. Meta-analysis and multidisciplinary consensus statement: exome sequencing is a first-tier clinical diagnostic test for individuals with neurodevelopmental disorders. Genet Med. 2019;21(11):2413–2421. PMCID: PMC6831729

2. Scocchia A, Wigby KM, Masser-Frye D, Del Campo M, Galarreta CI, Thorpe E, McEachern J, Robinson K, Gross A, Ajay SS, Rajan V, Perry DL, Belmont JW, Bentley DR, Jones MC, Taft RJ. Clinical whole genome sequencing as a first-tier test at a resource-limited dysmorphology clinic in Mexico. npj Genomic Med. 2019 Feb 14;4(1):1–12. PMCID: PMC6375919

3. Manickam K, McClain MR, Demmer LA, Biswas S, Kearney HM, Malinowski J, Massingham LJ, Miller D, Yu TW, Hisama FM. Exome and genome sequencing for pediatric patients with congenital anomalies or intellectual disability: an evidence-based clinical guideline of the American College of Medical Genetics and Genomics (ACMG). Genetics in Medicine. 2021 Nov 1;23(11):2029–2037. PMID: 34211152

4. Yaron Y, Ofen Glassner V, Mory A, Zunz Henig N, Kurolap A, Bar Shira A, Brabbing Goldstein D, Marom D, Ben Sira L, Baris Feldman H, Malinger G, Krajden Haratz K, Reches A. Exome sequencing as first-tier test for fetuses with severe central nervous system structural anomalies. Ultrasound in Obstetrics & Gynecology. 2022;60(1):59–67. PMCID: PMC9328397

5. van der Sanden BPGH, Schobers G, Corominas Galbany J, Koolen DA, Sinnema M, van Reeuwijk J, Stumpel CTRM, Kleefstra T, de Vries BBA, Ruiterkamp-Versteeg M, Leijsten N, Kwint M, Derks R, Swinkels H, den Ouden A, Pfundt R, Rinne T, de Leeuw N, Stegmann AP, Stevens SJ, van den Wijngaard A, Brunner HG, Yntema HG, Gilissen C, Nelen MR, Vissers LELM. The performance of genome sequencing as a first-tier test for neurodevelopmental disorders. Eur J Hum Genet. 2023 Jan;31(1):81–88. PMCID: PMC9822884

6. Cirillo L, Becherucci F. The evolving role of first-tier exome sequencing in medical diagnostics. Nephrol Dial Transplant. 2024 Mar 27;39(4):560–563. PMID: 37858299

7. Bodian DL, Klein E, Iyer RK, Wong WSW, Kothiyal P, Stauffer D, Huddleston KC, Gaither AD, Remsburg I, Khromykh A, Baker RL, Maxwell GL, Vockley JG, Niederhuber JE, Solomon BD. Utility of whole-genome sequencing for detection of newborn screening disorders in a population cohort of 1,696 neonates. Genet Med. 2016 Mar;18(3):221–230. PMID: 26334177

8. Bailey DB, Gehtland LM, Lewis MA, Peay H, Raspa M, Shone SM, Taylor JL, Wheeler AC, Cotten M, King NMP, Powell CM, Biesecker B, Bishop CE, Boyea BL, Duparc M, Harper BA, Kemper AR, Lee SN, Moultrie R, Okoniewski KC, Paquin RS, Pettit D, Porter KA, Zimmerman SJ. Early Check: translational science at the intersection of public health and newborn screening. BMC Pediatr. 2019 Jul 17;19(1):238. PMCID: PMC6636013

9. Foss KS, O’Daniel JM, Berg JS, Powell SN, Cadigan RJ, Kuczynski KJ, Milko LV, Saylor KW, Roberts M, Weck K, Henderson GE. The Rise of Population Genomic Screening: Characteristics of Current Programs and the Need for Evidence Regarding Optimal Implementation. Journal of Personalized Medicine. 2022 May;12(5):692. PMCID: PMC9145687

10. Buchanan AH, Lester Kirchner H, Schwartz MLB, Kelly MA, Schmidlen T, Jones LK, Hallquist MLG, Rocha H, Betts M, Schwiter R, Butry L, Lazzeri AL, Frisbie LR, Rahm AK, Hao J, Willard HF, Martin CL, Ledbetter DH, Williams MS, Sturm AC. Clinical outcomes of a genomic screening program for actionable genetic conditions. Genet Med. 2020 Nov;22(11):1874–1882. PMCID: PMC7605431

11. Casalino S, Frangione E, Chung M, MacDonald G, Chowdhary S, Mighton C, Faghfoury H, Bombard Y, Strug L, Pugh TJ, Simpson J, Arnoldo S, Aujla N, Bearss E, Binnie A, Borgundvaag B, Chertkow H, Clausen M, Dagher M, Devine L, Di Iorio D, Friedman SM, Fung CYJ, Gingras AC, Goneau LW, Kaushik D, Khan Z, Lapadula E, Lu T, Mazzulli T, McGeer A, McLeod SL, Morgan G, Richardson D, Singh H, Stern S, Taher A, Wong I, Zarei N, Greenfeld E, Hao L, Lebo M, Lane W, Noor A, Taher J, Lerner-Ellis J. Genome screening, reporting, and genetic counseling for healthy populations. Hum Genet. 2023 Feb;142(2):181–192. PMCID: PMC9638226

12. Chen T, Fan C, Huang Y, Feng J, Zhang Y, Miao J, Wang X, Li Y, Huang C, Jin W, Tang C, Feng L, Yin Y, Zhu B, Sun M, Liu X, Xiang J, Tan M, Jia L, Chen L, Huang H, Peng H, Sun X, Gu X, Peng Z, Zhu B, Zou H, Han L. Genomic Sequencing as a First-Tier Screening Test and Outcomes of Newborn Screening. JAMA Netw Open. 2023 Sep 5;6(9):e2331162. PMCID: PMC10474521

13. Green RC, Shah N, Genetti CA, Yu T, Zettler B, Uveges MK, Ceyhan-Birsoy O, Lebo MS, Pereira S, Agrawal PB, Parad RB, McGuire AL, Christensen KD, Schwartz TS, Rehm HL, Holm IA, Beggs AH, BabySeq Project Team. Actionability of unanticipated monogenic disease risks in newborn genomic screening: Findings from the BabySeq Project. Am J Hum Genet. 2023 Jul 6;110(7):1034–1045. PMCID: PMC10357495

14. Stone K. The Generation Study — Knowledge Hub [Internet]. GeNotes. [cited 2024 Aug 20]. Available from: https://www.genomicseducation.hee.nhs.uk/genotes/knowledge-hub/the-generation-study/

15. Stark Z, Scott RH. Genomic newborn screening for rare diseases. Nat Rev Genet. 2023 Nov;24(11):755–766. PMID: 37386126

16. Chung WK, Kanne SM, Hu Z. An Opportunity to Fill a Gap for Newborn Screening of Neurodevelopmental Disorders. Int J Neonatal Screen. 2024 Apr 16;10(2):33. PMCID: PMC11036277

17. Baple EL, Scott RH, Banka S, Buchanan J, Fish L, Wynn S, Wilkinson D, Ellard S, MacArthur DG, Stark Z. Exploring the benefits, harms and costs of genomic newborn screening for rare diseases. Nat Med. 2024 Jul;30(7):1823–1825. PMID: 38898121

18. Bowling KM, Thompson ML, Kelly MA, Scollon S, Slavotinek AM, Powell BC, Kirmse BM, Hendon LG, Brothers KB, Korf BR, Cooper GM, Greally JM, Hurst ACE. Return of non-ACMG recommended incidental genetic findings to pediatric patients: considerations and opportunities from experiences in genomic sequencing. Genome Medicine. 2022 Nov 21;14(1):131.

19. Woerner AC, Gallagher RC, Vockley J, Adhikari AN. The Use of Whole Genome and Exome Sequencing for Newborn Screening: Challenges and Opportunities for Population Health. Front Pediatr. 2021;9:663752. PMCID: PMC8326411

20. Horton R, Wright CF, Firth HV, Turnbull C, Lachmann R, Houlston RS, Lucassen A. Challenges of using whole genome sequencing in population newborn screening. BMJ. British Medical Journal Publishing Group; 2024 Mar 5;384:e077060. PMID: 38443063

21. Turnbull C, Firth HV, Wilkie AOM, Newman W, Raymond FL, Tomlinson I, Lachmann R, Wright CF, Wordsworth S, George A, McCartney M, Lucassen A. Population screening requires robust evidence—genomics is no exception. The Lancet. Elsevier; 2024 Feb 10;403(10426):583–586. PMID: 38070525

22. Zhao S, Agafonov O, Azab A, Stokowy T, Hovig E. Accuracy and efficiency of germline variant calling pipelines for human genome data. Sci Rep. 2020 Nov 19;10(1):20222. PMCID: PMC7678823

23. Olson ND, Wagner J, McDaniel J, Stephens SH, Westreich ST, Prasanna AG, Johanson E, Boja E, Maier EJ, Serang O, Jáspez D, Lorenzo-Salazar JM, Muñoz-Barrera A, Rubio-Rodríguez LA, Flores C, Kyriakidis K, Malousi A, Shafin K, Pesout T, Jain M, Paten B, Chang PC, Kolesnikov A, Nattestad M, Baid G, Goel S, Yang H, Carroll A, Eveleigh R, Bourgey M, Bourque G, Li G, Ma C, Tang L, Du Y, Zhang S, Morata J, Tonda R, Parra G, Trotta JR, Brueffer C, Demirkaya-Budak S, Kabakci-Zorlu D, Turgut D, Kalay Ö, Budak G, Narci K, Arslan E, Brown R, Johnson IJ, Dolgoborodov A, Semenyuk V, Jain A, Tetikol HS, Jain V, Ruehle M, Lajoie B, Roddey C, Catreux S, Mehio R, Ahsan MU, Liu Q, Wang K, Ebrahim Sahraeian SM, Fang LT, Mohiyuddin M, Hung C, Jain C, Feng H, Li Z, Chen L, Sedlazeck FJ, Zook JM. PrecisionFDA Truth Challenge V2: Calling variants from short and long reads in difficult-to-map regions. Cell Genom. 2022 Apr 27;2(5):100129. PMCID: PMC9205427

24. Fahed AC, Wang M, Homburger JR, Patel AP, Bick AG, Neben CL, Lai C, Brockman D, Philippakis A, Ellinor PT, Cassa CA, Lebo M, Ng K, Lander ES, Zhou AY, Kathiresan S, Khera AV. Polygenic background modifies penetrance of monogenic variants for tier 1 genomic conditions. Nat Commun. 2020 Aug 20;11(1):3635. PMCID: PMC7441381

25. Blair DR, Hoffmann TJ, Shieh JT. Common genetic variation associated with Mendelian disease severity revealed through cryptic phenotype analysis [Internet]. 2021 Aug p. 2021.08.26.21262300. Available from: https://www.medrxiv.org/content/10.1101/2021.08.26.21262300v1

26. Kingdom R, Beaumont RN, Wood AR, Weedon MN, Wright CF. Genetic modifiers of rare variants in monogenic developmental disorder loci. Nat Genet. 2024 May;56(5):861–868. PMCID: PMC11096126

27. Tukker AM, Royal CD, Bowman AB, McAllister KA. The Impact of Environmental Factors on Monogenic Mendelian Diseases. Toxicol Sci. 2021 Mar 2;181(1):3–12. PMCID: PMC8599782

28. Richards S, Aziz N, Bale S, Bick D, Das S, Gastier-Foster J, Grody WW, Hegde M, Lyon E, Spector E, Voelkerding K, Rehm HL. Standards and Guidelines for the Interpretation of Sequence Variants: A Joint Consensus Recommendation of the American College of Medical Genetics and Genomics and the Association for Molecular Pathology. Genet Med. 2015 May;17(5):405–424. PMCID: PMC4544753

29. Kingdom R, Wright CF. Incomplete Penetrance and Variable Expressivity: From Clinical Studies to Population Cohorts. Front Genet. 2022;13:920390. PMCID: PMC9380816

30. Risch NJ, Bressman SB, Senthil G, Ozelius LJ. Intragenic Cis and Trans Modification of Genetic Susceptibility in DYT1 Torsion Dystonia. Am J Hum Genet. 2007 Jun;80(6):1188–1193. PMCID: PMC1867106

31. Chen S, Parmigiani G. Meta-Analysis of BRCA1 and BRCA2 Penetrance. J Clin Oncol. 2007 Apr 10;25(11):1329–1333. PMCID: PMC2267287

32. Hoffmann TJ, Sakoda LC, Shen L, Jorgenson E, Habel LA, Liu J, Kvale MN, Asgari MM, Banda Y, Corley D, Kushi LH, Quesenberry CP, Schaefer C, Van Den Eeden SK, Risch N, Witte JS. Imputation of the Rare HOXB13 G84E Mutation and Cancer Risk in a Large Population-Based Cohort. PLoS Genet. 2015 Jan 28;11(1):e1004930. PMCID: PMC4309593

33. Menozzi E, Schapira AHV. Exploring the Genotype–Phenotype Correlation in GBA-Parkinson Disease: Clinical Aspects, Biomarkers, and Potential Modifiers. Front Neurol [Internet]. Frontiers; 2021 Jun 24 [cited 2024 Aug 28];12. Available from: https://www.frontiersin.org/journals/neurology/articles/10.3389/fneur.2021.694764/full

34. Roden DM, Pulley JM, Basford MA, Bernard GR, Clayton EW, Balser JR, Masys DR. Development of a large-scale de-identified DNA biobank to enable personalized medicine. Clin Pharmacol Ther. 2008 Sep;84(3):362–369. PMCID: PMC3763939

35. Kvale MN, Hesselson S, Hoffmann TJ, Cao Y, Chan D, Connell S, Croen LA, Dispensa BP, Eshragh J, Finn A, Gollub J, Iribarren C, Jorgenson E, Kushi LH, Lao R, Lu Y, Ludwig D, Mathauda GK, McGuire WB, Mei G, Miles S, Mittman M, Patil M, Quesenberry CP Jr, Ranatunga D, Rowell S, Sadler M, Sakoda LC, Shapero M, Shen L, Shenoy T, Smethurst D, Somkin CP, Van Den Eeden SK, Walter L, Wan E, Webster T, Whitmer RA, Wong S, Zau C, Zhan Y, Schaefer C, Kwok PY, Risch N. Genotyping Informatics and Quality Control for 100,000 Subjects in the Genetic Epidemiology Research on Adult Health and Aging (GERA) Cohort. Genetics. 2015 Aug 1;200(4):1051–1060. PMCID: PMC4574249

36. Carey DJ, Fetterolf SN, Davis FD, Faucett WA, Kirchner HL, Mirshahi U, Murray MF, Smelser DT, Gerhard GS, Ledbetter DH. The Geisinger MyCode community health initiative: an electronic health record-linked biobank for precision medicine research. Genet Med. 2016;18(9):906–913. PMCID: PMC4981567

37. Bycroft C, Freeman C, Petkova D, Band G, Elliott LT, Sharp K, Motyer A, Vukcevic D, Delaneau O, O’Connell J, Cortes A, Welsh S, Young A, Effingham M, McVean G, Leslie S, Allen N, Donnelly P, Marchini J. The UK Biobank resource with deep phenotyping and genomic data. Nature. 2018 Oct;562(7726):203–209. PMCID: PMC6786975

38. Belbin GM, Cullina S, Wenric S, Soper ER, Glicksberg BS, Torre D, Moscati A, Wojcik GL, Shemirani R, Beckmann ND, Cohain A, Sorokin EP, Park DS, Ambite JL, Ellis S, Auton A, Bottinger EP, Cho JH, Loos RJF, Abul-Husn NS, Zaitlen NA, Gignoux CR, Kenny EE. Toward a fine-scale population health monitoring system. Cell. Elsevier; 2021 Apr 15;184(8):2068–2083.e11. PMID: 33861964

39. Kurki MI, Karjalainen J, Palta P, Sipilä TP, Kristiansson K, Donner KM, Reeve MP, Laivuori H, Aavikko M, Kaunisto MA, Loukola A, Lahtela E, Mattsson H, Laiho P, Della Briotta Parolo P, Lehisto AA, Kanai M, Mars N, Rämö J, Kiiskinen T, Heyne HO, Veerapen K, Rüeger S, Lemmelä S, Zhou W, Ruotsalainen S, Pärn K, Hiekkalinna T, Koskelainen S, Paajanen T, Llorens V, Gracia-Tabuenca J, Siirtola H, Reis K, Elnahas AG, Sun B, Foley CN, Aalto-Setälä K, Alasoo K, Arvas M, Auro K, Biswas S, Bizaki-Vallaskangas A, Carpen O, Chen CY, Dada OA, Ding Z, Ehm MG, Eklund K, Färkkilä M, Finucane H, Ganna A, Ghazal A, Graham RR, Green EM, Hakanen A, Hautalahti M, Hedman ÅK, Hiltunen M, Hinttala R, Hovatta I, Hu X, Huertas-Vazquez A, Huilaja L, Hunkapiller J, Jacob H, Jensen JN, Joensuu H, John S, Julkunen V, Jung M, Junttila J, Kaarniranta K, Kähönen M, Kajanne R, Kallio L, Kälviäinen R, Kaprio J, Kerimov N, Kettunen J, Kilpeläinen E, Kilpi T, Klinger K, Kosma VM, Kuopio T, Kurra V, Laisk T, Laukkanen J, Lawless N, Liu A, Longerich S, Mägi R, Mäkelä J, Mäkitie A, Malarstig A, Mannermaa A, Maranville J, Matakidou A, Meretoja T, Mozaffari SV, Niemi MEK, Niemi M, Niiranen T, O’Donnell CJ, Obeidat M, Okafo G, Ollila HM, Palomäki A, Palotie T, Partanen J, Paul DS, Pelkonen M, Pendergrass RK, Petrovski S, Pitkäranta A, Platt A, Pulford D, Punkka E, Pussinen P, Raghavan N, Rahimov F, Rajpal D, Renaud NA, Riley-Gillis B, Rodosthenous R, Saarentaus E, Salminen A, Salminen E, Salomaa V, Schleutker J, Serpi R, Shen H yi, Siegel R, Silander K, Siltanen S, Soini S, Soininen H, Sul JH, Tachmazidou I, Tasanen K, Tienari P, Toppila-Salmi S, Tukiainen T, Tuomi T, Turunen JA, Ulirsch JC, Vaura F, Virolainen P, Waring J, Waterworth D, Yang R, Nelis M, Reigo A, Metspalu A, Milani L, Esko T, Fox C, Havulinna AS, Perola M, Ripatti S, Jalanko A, Laitinen T, Mäkelä TP, Plenge R, McCarthy M, Runz H, Daly MJ, Palotie A. FinnGen provides genetic insights from a well-phenotyped isolated population. Nature. 2023;613(7944):508–518. PMCID: PMC9849126

40. Johnson R, Ding Y, Bhattacharya A, Knyazev S, Chiu A, Lajonchere C, Geschwind DH, Pasaniuc B. The UCLA ATLAS Community Health Initiative: Promoting precision health research in a diverse biobank. Cell Genom. 2023 Jan 11;3(1):100243. PMCID: PMC9903668

41. All of Us Research Program Genomics Investigators. Genomic data in the All of Us Research Program. Nature. 2024 Mar;627(8003):340–346. PMCID: PMC10937371

42. Wright CF, Sharp LN, Jackson L, Murray A, Ware JS, MacArthur DG, Rehm HL, Patel KA, Weedon MN. Guidance for estimating penetrance of monogenic disease-causing variants in population cohorts. Nat Genet. 2024 Jul 29; PMID: 39075210

43. Wright CF, West B, Tuke M, Jones SE, Patel K, Laver TW, Beaumont RN, Tyrrell J, Wood AR, Frayling TM, Hattersley AT, Weedon MN. Assessing the Pathogenicity, Penetrance, and Expressivity of Putative Disease-Causing Variants in a Population Setting. Am J Hum Genet. 2019 Feb 7;104(2):275–286. PMCID: PMC6369448

44. Forrest IS, Chaudhary K, Vy HMT, Petrazzini BO, Bafna S, Jordan DM, Rocheleau G, Loos RJF, Nadkarni GN, Cho JH, Do R. Population-Based Penetrance of Deleterious Clinical Variants. JAMA. 2022 Jan 25;327(4):350–359. PMCID: PMC8790667

45. Mirshahi UL, Colclough K, Wright CF, Wood AR, Beaumont RN, Tyrrell J, Laver TW, Stahl R, Golden A, Goehringer JM, Frayling TF, Hattersley AT, Carey DJ, Weedon MN, Patel KA. Reduced penetrance of MODY-associated HNF1A/HNF4A variants but not GCK variants in clinically unselected cohorts. Am J Hum Genet. 2022 Nov 3;109(11):2018–2028. PMCID: PMC9674944

46. Bastarache L, Peterson JF. Penetrance of Deleterious Clinical Variants. JAMA. 2022 May 17;327(19):1926–1927. PMCID: PMC9350877

47. Landrum MJ, Lee JM, Benson M, Brown GR, Chao C, Chitipiralla S, Gu B, Hart J, Hoffman D, Jang W, Karapetyan K, Katz K, Liu C, Maddipatla Z, Malheiro A, McDaniel K, Ovetsky M, Riley G, Zhou G, Holmes JB, Kattman BL, Maglott DR. ClinVar: improving access to variant interpretations and supporting evidence. Nucleic Acids Res. 2018 Jan 4;46(D1):D1062–D1067. PMCID: PMC5753237

48. Rehm HL, Berg JS, Brooks LD, Bustamante CD, Evans JP, Landrum MJ, Ledbetter DH, Maglott DR, Martin CL, Nussbaum RL, Plon SE, Ramos EM, Sherry ST, Watson MS, ClinGen. ClinGen--the Clinical Genome Resource. N Engl J Med. 2015 Jun 4;372(23):2235–2242. PMCID: PMC4474187

49. Hamosh A, Scott AF, Amberger JS, Bocchini CA, McKusick VA. Online Mendelian Inheritance in Man (OMIM), a knowledgebase of human genes and genetic disorders. Nucleic Acids Res. 2005 Jan 1;33(Database issue):D514–517. PMCID: PMC539987

50. Sudlow C, Gallacher J, Allen N, Beral V, Burton P, Danesh J, Downey P, Elliott P, Green J, Landray M, Liu B, Matthews P, Ong G, Pell J, Silman A, Young A, Sprosen T, Peakman T, Collins R. UK Biobank: An Open Access Resource for Identifying the Causes of a Wide Range of Complex Diseases of Middle and Old Age. PLoS Med. 2015 Mar 31;12(3):e1001779. PMCID: PMC4380465

51. The “All of Us” Research Program. New England Journal of Medicine. Massachusetts Medical Society; 2019 Aug 15;381(7):668–676. PMID: 31412182

52. MacArthur DG, Balasubramanian S, Frankish A, Huang N, Morris J, Walter K, Jostins L, Habegger L, Pickrell JK, Montgomery SB, Albers CA, Zhang Z, Conrad DF, Lunter G, Zheng H, Ayub Q, DePristo MA, Banks E, Hu M, Handsaker RE, Rosenfeld J, Fromer M, Jin M, Mu XJ, Khurana E, Ye K, Kay M, Saunders GI, Suner MM, Hunt T, Barnes IHA, Amid C, Carvalho-Silva DR, Bignell AH, Snow C, Yngvadottir B, Bumpstead S, Cooper DN, Xue Y, Romero IG, Wang J, Li Y, Gibbs RA, McCarroll SA, Dermitzakis ET, Pritchard JK, Barrett JC, Harrow J, Hurles ME, Gerstein MB, Tyler-Smith C. A systematic survey of loss-of-function variants in human protein-coding genes. Science. 2012 Feb 17;335(6070):823–828. PMCID: PMC3299548

53. Karczewski KJ, Francioli LC, Tiao G, Cummings BB, Alföldi J, Wang Q, Collins RL, Laricchia KM, Ganna A, Birnbaum DP, Gauthier LD, Brand H, Solomonson M, Watts NA, Rhodes D, Singer-Berk M, England EM, Seaby EG, Kosmicki JA, Walters RK, Tashman K, Farjoun Y, Banks E, Poterba T, Wang A, Seed C, Whiffin N, Chong JX, Samocha KE, Pierce-Hoffman E, Zappala Z, O’Donnell-Luria AH, Minikel EV, Weisburd B, Lek M, Ware JS, Vittal C, Armean IM, Bergelson L, Cibulskis K, Connolly KM, Covarrubias M, Donnelly S, Ferriera S, Gabriel S, Gentry J, Gupta N, Jeandet T, Kaplan D, Llanwarne C, Munshi R, Novod S, Petrillo N, Roazen D, Ruano-Rubio V, Saltzman A, Schleicher M, Soto J, Tibbetts K, Tolonen C, Wade G, Talkowski ME, Neale BM, Daly MJ, MacArthur DG. The mutational constraint spectrum quantified from variation in 141,456 humans. Nature. 2020 May;581(7809):434–443. PMCID: PMC7334197

54. Singer-Berk M, Gudmundsson S, Baxter S, Seaby EG, England E, Wood JC, Son RG, Watts NA, Karczewski KJ, Harrison SM, MacArthur DG, Rehm HL, O’Donnell-Luria A. Advanced variant classification framework reduces the false positive rate of predicted loss-of-function variants in population sequencing data. Am J Hum Genet. 2023 Sep 7;110(9):1496–1508. PMCID: PMC10502856

55. Gudmundsson S, Singer-Berk M, Stenton SL, Goodrich JK, Wilson MW, Einson J, Watts NA, Lappalainen T, Rehm HL, MacArthur DG, O’Donnell-Luria A. Exploring penetrance of clinically relevant variants in over 800,000 humans from the Genome Aggregation Database. bioRxiv. 2024 Jun 13;2024.06.12.593113. PMCID: PMC11195293

56. Morales J, Pujar S, Loveland JE, Astashyn A, Bennett R, Berry A, Cox E, Davidson C, Ermolaeva O, Farrell CM, Fatima R, Gil L, Goldfarb T, Gonzalez JM, Haddad D, Hardy M, Hunt T, Jackson J, Joardar VS, Kay M, Kodali VK, McGarvey KM, McMahon A, Mudge JM, Murphy DN, Murphy MR, Rajput B, Rangwala SH, Riddick LD, Thibaud-Nissen F, Threadgold G, Vatsan AR, Wallin C, Webb D, Flicek P, Birney E, Pruitt KD, Frankish A, Cunningham F, Murphy TD. A joint NCBI and EMBL-EBI transcript set for clinical genomics and research. Nature. 2022 Apr;604(7905):310–315. PMCID: PMC9007741

57. Forrest IS, Duffy Á, Park JK, Vy HMT, Pasquale LR, Nadkarni GN, Cho JH, Do R. Genome-first evaluation with exome sequence and clinical data uncovers underdiagnosed genetic disorders in a large healthcare system. Cell Rep Med. 2024 Apr 19;5(5):101518. PMCID: PMC11148562

58. Bastarache L, Hughey JJ, Hebbring S, Marlo J, Zhao W, Ho WT, Van Driest SL, McGregor TL, Mosley JD, Wells QS, Temple M, Ramirez AH, Carroll R, Osterman T, Edwards T, Ruderfer D, Velez Edwards DR, Hamid R, Cogan J, Glazer A, Wei WQ, Feng Q, Brilliant M, Zhao ZJ, Cox NJ, Roden DM, Denny JC. Phenotype risk scores identify patients with unrecognized Mendelian disease patterns. Science. 2018 16;359(6381):1233–1239. PMCID: PMC5959723

59. Bastarache L, Hughey JJ, Goldstein JA, Bastraache JA, Das S, Zaki NC, Zeng C, Tang LA, Roden DM, Denny JC. Improving the phenotype risk score as a scalable approach to identifying patients with Mendelian disease. Journal of the American Medical Informatics Association. 2019 Dec 1;26(12):1437–1447.

60. Lindeboom RGH, Vermeulen M, Lehner B, Supek F. The impact of nonsense-mediated mRNA decay on genetic disease, gene editing and cancer immunotherapy. Nat Genet. 2019 Nov;51(11):1645–1651. PMCID: PMC6858879

61. Dyle MC, Kolakada D, Cortazar MA, Jagannathan S. How to get away with nonsense: Mechanisms and consequences of escape from nonsense-mediated RNA decay. Wiley Interdiscip Rev RNA. 2020 Jan;11(1):e1560. PMCID: PMC10685860

62. Johnson AF, Nguyen HT, Veitia RA. Causes and effects of haploinsufficiency. Biol Rev Camb Philos Soc. 2019 Oct;94(5):1774–1785. PMID: 31149781

63. Breiman L. Random Forests. Machine Learning. 2001 Oct 1;45(1):5–32.

64. sklearn.metrics.average_precision_score — scikit-learn 0.23.1 documentation [Internet]. [cited 2020 Jul 2]. Available from: https://scikit-learn.org/stable/modules/generated/sklearn.metrics.average_precision_score.html

65. Lundberg SM, Lee SI. A Unified Approach to Interpreting Model Predictions. Advances in Neural Information Processing Systems [Internet]. Curran Associates, Inc.; 2017 [cited 2025 Jan 31]. Available from: https://papers.nips.cc/paper_files/paper/2017/hash/8a20a8621978632d76c43dfd28b67767-Abstract.html

66. Schubach M, Maass T, Nazaretyan L, Röner S, Kircher M. CADD v1.7: using protein language models, regulatory CNNs and other nucleotide-level scores to improve genome-wide variant predictions. Nucleic Acids Research. 2024 Jan 5;52(D1):D1143–D1154. PMCID: PMC10767851

67. Beaumont RN, Hawkes G, Gunning AC, Wright CF. Clustering of predicted loss-of-function variants in genes linked with monogenic disease can explain incomplete penetrance. Genome Medicine. 2024 Apr 26;16(1):64. PMCID: PMC11046769

68. Chung CCY, Hue SPY, Ng NYT, Doong PHL, Hong Kong Genome Project, Chu ATW, Chung BHY. Meta-analysis of the diagnostic and clinical utility of exome and genome sequencing in pediatric and adult patients with rare diseases across diverse populations. Genet Med. 2023 Sep;25(9):100896. PMID: 37191093

69. Amendola LM, Jarvik GP, Leo MC, McLaughlin HM, Akkari Y, Amaral MD, Berg JS, Biswas S, Bowling KM, Conlin LK, Cooper GM, Dorschner MO, Dulik MC, Ghazani AA, Ghosh R, Green RC, Hart R, Horton C, Johnston JJ, Lebo MS, Milosavljevic A, Ou J, Pak CM, Patel RY, Punj S, Richards CS, Salama J, Strande NT, Yang Y, Plon SE, Biesecker LG, Rehm HL. Performance of ACMG-AMP Variant-Interpretation Guidelines among Nine Laboratories in the Clinical Sequencing Exploratory Research Consortium. Am J Hum Genet. 2016 Jun 2;98(6):1067–1076. PMCID: PMC4908185

70. Jaganathan K, Kyriazopoulou Panagiotopoulou S, McRae JF, Darbandi SF, Knowles D, Li YI, Kosmicki JA, Arbelaez J, Cui W, Schwartz GB, Chow ED, Kanterakis E, Gao H, Kia A, Batzoglou S, Sanders SJ, Farh KKH. Predicting Splicing from Primary Sequence with Deep Learning. Cell. 2019 Jan 24;176(3):535–548.e24. PMID: 30661751

71. Kingsmore SF, Smith LD, Kunard CM, Bainbridge M, Batalov S, Benson W, Blincow E, Caylor S, Chambers C, Del Angel G, Dimmock DP, Ding Y, Ellsworth K, Feigenbaum A, Frise E, Green RC, Guidugli L, Hall KP, Hansen C, Hobbs CA, Kahn SD, Kiel M, Van Der Kraan L, Krilow C, Kwon YH, Madhavrao L, Le J, Lefebvre S, Mardach R, Mowrey WR, Oh D, Owen MJ, Powley G, Scharer G, Shelnutt S, Tokita M, Mehtalia SS, Oriol A, Papadopoulos S, Perry J, Rosales E, Sanford E, Schwartz S, Tran D, Reese MG, Wright M, Veeraraghavan N, Wigby K, Willis MJ, Wolen AR, Defay. T. A genome sequencing system for universal newborn screening, diagnosis, and precision medicine for severe genetic diseases. Am J Hum Genet. 2022 Sep 1;109(9):1605–1619. PMCID: PMC9502059

72. Adhikari AN, Gallagher RC, Wang Y, Currier RJ, Amatuni G, Bassaganyas L, Chen F, Kundu K, Kvale M, Mooney SD, Nussbaum RL, Randi SS, Sanford J, Shieh JT, Srinivasan R, Sunderam U, Tang H, Vaka D, Zou Y, Koenig BA, Kwok PY, Risch N, Puck JM, Brenner SE. The Role of Exome Sequencing in Newborn Screening for Inborn Errors of Metabolism. Nat Med. 2020 Sep;26(9):1392–1397. PMCID: PMC8800147

73. Reich C, Ostropolets A, Ryan P, Rijnbeek P, Schuemie M, Davydov A, Dymshyts D, Hripcsak G. OHDSI Standardized Vocabularies-a large-scale centralized reference ontology for international data harmonization. J Am Med Inform Assoc. 2024 Feb 16;31(3):583–590. PMCID: PMC10873827

74. OMOP Common Data Model [Internet]. [cited 2024 Aug 20]. Available from: https://ohdsi.github.io/CommonDataModel/

75. Köhler S, Gargano M, Matentzoglu N, Carmody LC, Lewis-Smith D, Vasilevsky NA, Danis D, Balagura G, Baynam G, Brower AM, Callahan TJ, Chute CG, Est JL, Galer PD, Ganesan S, Griese M, Haimel M, Pazmandi J, Hanauer M, Harris NL, Hartnett MJ, Hastreiter M, Hauck F, He Y, Jeske T, Kearney H, Kindle G, Klein C, Knoflach K, Krause R, Lagorce D, McMurry JA, Miller JA, Munoz-Torres MC, Peters RL, Rapp CK, Rath AM, Rind SA, Rosenberg AZ, Segal MM, Seidel MG, Smedley D, Talmy T, Thomas Y, Wiafe SA, Xian J, Yüksel Z, Helbig I, Mungall CJ, Haendel MA, Robinson PN. The Human Phenotype Ontology in 2021. Nucleic Acids Res. 2021 Jan 8;49(D1):D1207–D1217. PMCID: PMC7778952

76. Schriml LM, Mitraka E, Munro J, Tauber B, Schor M, Nickle L, Felix V, Jeng L, Bearer C, Lichenstein R, Bisordi K, Campion N, Hyman B, Kurland D, Oates CP, Kibbey S, Sreekumar P, Le C, Giglio M, Greene C. Human Disease Ontology 2018 update: classification, content and workflow expansion. Nucleic Acids Res. 2019 08;47(D1):D955–D962. PMCID: PMC6323977

77. Orphanet [Internet]. [cited 2024 Aug 20]. Available from: https://www.orpha.net/

78. Köhler S, Carmody L, Vasilevsky N, Jacobsen JOB, Danis D, Gourdine JP, Gargano M, Harris NL, Matentzoglu N, McMurry JA, Osumi-Sutherland D, Cipriani V, Balhoff JP, Conlin T, Blau H, Baynam G, Palmer R, Gratian D, Dawkins H, Segal M, Jansen AC, Muaz A, Chang WH, Bergerson J, Laulederkind SJF, Yüksel Z, Beltran S, Freeman AF, Sergouniotis PI, Durkin D, Storm AL, Hanauer M, Brudno M, Bello SM, Sincan M, Rageth K, Wheeler MT, Oegema R, Lourghi H, Della Rocca MG, Thompson R, Castellanos F, Priest J, Cunningham-Rundles C, Hegde A, Lovering RC, Hajek C, Olry A, Notarangelo L, Similuk M, Zhang XA, Gómez-Andrés D, Lochmüller H, Dollfus H, Rosenzweig S, Marwaha S, Rath A, Sullivan K, Smith C, Milner JD, Leroux D, Boerkoel CF, Klion A, Carter MC, Groza T, Smedley D, Haendel MA, Mungall C, Robinson PN. Expansion of the Human Phenotype Ontology (HPO) knowledge base and resources. Nucleic Acids Res. 2019 Jan 8;47(Database issue):D1018–D1027. PMCID: PMC6324074

79. Martin FJ, Amode MR, Aneja A, Austine-Orimoloye O, Azov AG, Barnes I, Becker A, Bennett R, Berry A, Bhai J, Bhurji SK, Bignell A, Boddu S, Branco Lins PR, Brooks L, Ramaraju SB, Charkhchi M, Cockburn A, Da Rin Fiorretto L, Davidson C, Dodiya K, Donaldson S, El Houdaigui B, El Naboulsi T, Fatima R, Giron CG, Genez T, Ghattaoraya GS, Martinez JG, Guijarro C, Hardy M, Hollis Z, Hourlier T, Hunt T, Kay M, Kaykala V, Le T, Lemos D, Marques-Coelho D, Marugán JC, Merino GA, Mirabueno LP, Mushtaq A, Hossain SN, Ogeh DN, Sakthivel MP, Parker A, Perry M, Piližota I, Prosovetskaia I, Pérez-Silva JG, Salam AIA, Saraiva-Agostinho N, Schuilenburg H, Sheppard D, Sinha S, Sipos B, Stark W, Steed E, Sukumaran R, Sumathipala D, Suner MM, Surapaneni L, Sutinen K, Szpak M, Tricomi FF, Urbina-Gómez D, Veidenberg A, Walsh TA, Walts B, Wass E, Willhoft N, Allen J, Alvarez-Jarreta J, Chakiachvili M, Flint B, Giorgetti S, Haggerty L, Ilsley GR, Loveland JE, Moore B, Mudge JM, Tate J, Thybert D, Trevanion SJ, Winterbottom A, Frankish A, Hunt SE, Ruffier M, Cunningham F, Dyer S, Finn RD, Howe KL, Harrison PW, Yates AD, Flicek P. Ensembl 2023. Nucleic Acids Res. 2023 Jan 6;51(D1):D933–D941. PMCID: PMC9825606

80. pyensembl package — pyensembl 0.8.10 documentation [Internet]. [cited 2024 Aug 20]. Available from: https://pyensembl.readthedocs.io/en/latest/pyensembl.html

81. Luebbert L, Pachter L. Efficient querying of genomic reference databases with gget. Bioinformatics. 2023 Jan 1;39(1):btac836. PMCID: PMC9835474

82. Tan AL, Gonçalves RS, Yuan W, Brat GA, EHR TC for CC of C 19 by, Gentleman R, Kohane IS. Implications of mappings between ICD clinical diagnosis codes and Human Phenotype Ontology terms [Internet]. arXiv; 2024 [cited 2024 Aug 16]. Available from: http://arxiv.org/abs/2407.08874

83. SNOMED CT [Internet]. U.S. National Library of Medicine; [cited 2020 Jul 10]. Available from: https://www.nlm.nih.gov/healthit/snomedct/index.html

84. McArthur E, Bastarache L, Capra JA. Linking rare and common disease vocabularies by mapping between the human phenotype ontology and phecodes. JAMIA Open. 2023 Apr;6(1):ooad007. PMCID: PMC9976874

85. Bastarache L. Using Phecodes for Research with the Electronic Health Record: From PheWAS to PheRS. Annu Rev Biomed Data Sci. 2021 Jul 20;4:1–19. PMCID: PMC9307256

86. Schuyler PL, Hole WT, Tuttle MS, Sherertz DD. The UMLS Metathesaurus: representing different views of biomedical concepts. Bull Med Libr Assoc. 1993 Apr;81(2):217–222. PMCID: PMC225764

87. Pang C, Sollie A, Sijtsma A, Hendriksen D, Charbon B, de Haan M, de Boer T, Kelpin F, Jetten J, van der Velde JK, Smidt N, Sijmons R, Hillege H, Swertz MA. SORTA: a system for ontology-based re-coding and technical annotation of biomedical phenotype data. Database (Oxford). 2015;2015:bav089. PMCID: PMC4574036

88. Athena [Internet]. [cited 2024 Aug 20]. Available from: https://athena.ohdsi.org/search-terms/start

89. Pedregosa F, Varoquaux G, Gramfort A, Michel V, Thirion B, Grisel O, Blondel M, Prettenhofer P, Weiss R, Dubourg V, Vanderplas J, Passos A, Cournapeau D, Brucher M, Perrot M, Duchesnay É. Scikit-learn: Machine Learning in Python. Journal of Machine Learning Research. 2011;12(85):2825–2830.

90. Backman JD, Li AH, Marcketta A, Sun D, Mbatchou J, Kessler MD, Benner C, Liu D, Locke AE, Balasubramanian S, Yadav A, Banerjee N, Gillies CE, Damask A, Liu S, Bai X, Hawes A, Maxwell E, Gurski L, Watanabe K, Kosmicki JA, Rajagopal V, Mighty J, Jones M, Mitnaul L, Stahl E, Coppola G, Jorgenson E, Habegger L, Salerno WJ, Shuldiner AR, Lotta LA, Overton JD, Cantor MN, Reid JG, Yancopoulos G, Kang HM, Marchini J, Baras A, Abecasis GR, Ferreira MAR. Exome sequencing and analysis of 454,787 UK Biobank participants. Nature. 2021;599(7886):628–634. PMCID: PMC8596853

91. Danecek P, Bonfield JK, Liddle J, Marshall J, Ohan V, Pollard MO, Whitwham A, Keane T, McCarthy SA, Davies RM, Li H. Twelve years of SAMtools and BCFtools. GigaScience. 2021 Feb 1;10(2):giab008.

92. Hail Team. Hail 0.2. Available from: https://github.com/hail-is/hail

93. McLaren W, Gil L, Hunt SE, Riat HS, Ritchie GRS, Thormann A, Flicek P, Cunningham F. The Ensembl Variant Effect Predictor. Genome Biology. 2016 Jun 6;17(1):122. PMCID: PMC4893825

94. UK Biobank Whole Exome Sequencing 300k Release: Analysis Best Practices [Internet]. Available from: https://biobank.ndph.ox.ac.uk/showcase/ukb/docs/UKB_WES_AnalysisBestPractices.pdf

95. Wang X. Firth logistic regression for rare variant association tests. Front Genet [Internet]. Frontiers; 2014 [cited 2021 Aug 6];0. Available from: https://www.frontiersin.org/articles/10.3389/fgene.2014.00187/full

96. Seabold S, Perktold J. Statsmodels: Econometric and Statistical Modeling with Python. Proceedings of the 9th Python in Science Conference. 2010 Jan 1;2010.

97. Virtanen P, Gommers R, Oliphant TE, Haberland M, Reddy T, Cournapeau D, Burovski E, Peterson P, Weckesser W, Bright J, van der Walt SJ, Brett M, Wilson J, Millman KJ, Mayorov N, Nelson ARJ, Jones E, Kern R, Larson E, Carey CJ, Polat İ, Feng Y, Moore EW, VanderPlas J, Laxalde D, Perktold J, Cimrman R, Henriksen I, Quintero EA, Harris CR, Archibald AM, Ribeiro AH, Pedregosa F, van Mulbregt P. SciPy 1.0: fundamental algorithms for scientific computing in Python. Nat Methods. Nature Publishing Group; 2020 Mar;17(3):261–272.

98. Efron B, Tibshirani RJ. An Introduction to the Bootstrap. 1 edition. New York: Chapman and Hall/CRC; 1993.

99. pandas: powerful Python data analysis toolkit [Internet]. pandas; 2022 [cited 2022 Apr 18]. Available from: https://github.com/pandas-dev/pandas

100. Borenstein M, Hedges LV, Higgins JPT, Rothstein HR. A basic introduction to fixed-effect and random-effects models for meta-analysis. Res Synth Methods. 2010 Apr;1(2):97–111. PMID: 26061376

101. daverblair. daverblair/pLoFPenetranceScripts [Internet]. 2024 [cited 2024 Dec 5]. Available from: https://github.com/daverblair/pLoFPenetranceScripts

