## Supplementary Methods, Figures and Tables for "Reduced Penetrance is Common Among Predicted Loss-of-Function Variants and is Likely Driven by Residual Allelic Activity"

***Supplementary Information***

David R. Blair and Neil Risch

This document contains the Supplementary Methods, Figures, and Tables referenced in the main text.

**Supplementary Methods**

*Symptom-driven Expression Model Inference: the UK Biobank*

Symptom expression is not necessarily indicative of Mendelian *disease* expression, as some symptoms are quite common in the general population and may simply represent background noise. To identify pLoF carriers that have a high likelihood of disease expression, we developed the following probability model for symptom-driven disease expression (equivalent to Eqn. 7, Main Text):

$\begin{aligned} P\left( E_{i}=1 | \vec{S}_{i},\delta,\theta,\pi\right)=F\left[ \text{log}\left( \frac{\pi\times P\left( \vec{S}_{i} | \delta\right)}{\left( 1-\pi\right)\times P\left( \vec{S}_{i} | \theta\right)} \right) \right],\#\left( 1 \right) \end{aligned}$

where $E_{i}=1$denotes that the disease of interest was expressed in the *i*th carrier. The parameter $\pi$ is the prior probability of disease expression among all pathogenic variant carriers. $P\left( \vec{S}_{i} | \theta\right)$ indicates the probability that a set of symptoms $\vec{S}_{i}$ was expressed according to some general background distribution, and $P\left( \vec{S}_{i} | \delta\right)$ denotes the probability that this symptom set was instead expressed by an affected Mendelian disease variant carrier. The function $F[x]$ simply denotes the logistic function. An illustration of this model is depicted in Supplementary Figure 2A.

For this symptom-driven expression model to be effective, the Mendelian disease-specific symptom model (i.e. $P\left( \vec{S}_{i} | \delta\right)$), the background symptom model (i.e. $P\left( \vec{S}_{i} | \theta\right)$) and the expression prior probability (i.e. $\pi$) must be either known *a priori* or estimated from the data. Estimating $\pi$ from the data is relatively straightforward (see below). For the sake of simplicity, we used the background symptom expression models that we constructed for PheRS estimation to define $P\left( \vec{S}_{i} | \hat{\theta} \right)$ (see Eqn. 5 and 6, Main Text for details). Specifying and estimating the disease-specific expression models was more complex. In particular, the independence assumption invoked for PheRS estimation is very unlikely to hold for Mendelian diseases, as it is the co-occurrence of multiple symptoms that typically defines a Mendelian phenotype.

To overcome this issue, we assumed that $P\left( \vec{S}_{i} | \delta\right)$ follows an arbitrary distribution over symptoms sets. More specifically, let $\delta$ define a multinomial distribution over all possible expressed symptom sets (i.e. all possible sets except the empty set), such that the distribution is defined by a parameter set with cardinality $2^{K}-1$, where *K* denotes the total number of symptoms annotated to some disease of interest. Clearly, even for modest values of *K*, the dimensionality of the model becomes unwieldly, so we made the simplifying assumption that the possible set of symptoms was much smaller than $2^{K}-1$ (i.e. many of the multinomial distribution parameters are equal to 0). Practically, we assumed that only $M_{\text{Obs}}+1$ symptom sets were possible, where $M_{\text{Obs}}$denotes the number of unique symptom-sets observed among a collection of pathogenic variant carriers. The $+1$ term allows for the addition of a generic symptom set that accounts for any non-empty set that was not observed among these carriers; this allows the model to be applied to new datasets that may have symptom sets that were unobserved in the training data (see below). Note, the total complement of symptom sets observed among any collection of pLoF carriers was very sparse compared to the cardinality of all possible sets, such that $M_{\text{Obs}}+1$ was generally in the tens to hundreds rather than in the thousands to hundreds-of-thousands.

With this framework in place, the Mendelian disease symptom expression model was defined as:

$$P\left( \vec{S}_{i}=\vec{s} | \delta\right)=\sum_{m=1}^{M_{\text{Obs}}+1} \delta_{m}\times\boldsymbol{1(}\vec{s}\equiv\mathcal{S}_{\mathcal{m}})$$

where $\boldsymbol{1}(\vec{s}\equiv\mathcal{S}_{\mathcal{m}})$ is an indicator function that returns 1 if and only if the observed symptom set $\vec{s}$ is identical to the symptom set whose expression probability is defined by $\delta_{m}$ (denoted $\mathcal{S}_{\mathcal{m}}$ in the equation above). The symptom expression model defined in Eqn. 1 can then be used to specify the following likelihood for the observed symptom data:

$$\begin{aligned} P\left( \boldsymbol{S=s} | E,\theta,\delta,\pi\right)=\prod_{i=1}^{N} \sum_{E_{i}\in0,1} \left[ \pi\times P\left( \vec{S}_{i}=\vec{s} | \delta\right) \right]^{E_{i}}\times\left[ \left( 1-\pi\right)\times P\left( \vec{S}_{i}=\vec{s} | \theta\right) \right]^{1-E_{i}},\#\left( 2 \right) \end{aligned}$$

where $\boldsymbol{S}$ denotes the matrix of diagnosed symptoms across the *N* pLoF carriers. By maximizing this likelihood conditional on the symptom data observed among a set disease-specific pLoF carriers, estimates for the model parameters (specifically, $\pi$ and $\delta$) were obtained for each disease.

Practically, many of the diseases that we investigated had only a handful of pLoF carriers available for training. Therefore, we regularized the model parameter estimates by assuming that they were drawn from uniform Beta/Dirichlet distributions (for $\pi$ and $\delta$ respectively). The model specified in Eqn. 1 was thus fit by maximizing a lower-bound on the marginal likelihood using variational Bayesian inference^1^. The posterior distributions over the individual expression probabilities, denoted $P\left( E_{i}=1 | \vec{S}_{i},\hat{\delta},\hat{\theta},\hat{\pi} \right)$ for the *i*th carrier, were generated automatically during inference. In practice, we fit the disease-specific expression models only within the UKBB (given its larger sample size) using the SymptomSetModel software package^2^. Models were fit by first inferring a dataset-wide background distribution (using the using BuildBackgroundModel function) followed by directly optimizing the penetrance prior and disease-specific expression models (using the FitPenetranceModel function). The inferred model parameters (and subject-level expression probabilities, see Eqn. 1) were then written to disk, enabling the models to be applied to new datasets (i.e. the AoU Biobank).

*Symptom-driven Expression Model Validation: All of Us Research Program*

The symptom-driven expression model defined in Eqn. 1 is technically unsupervised, meaning that it does not require labeled training data for inference. Nevertheless, it remains at risk for overfitting just like any other model. Therefore, rather than independently fitting the model in each biobank, we used the AoU biobank as a validation dataset to ensure that our symptom-driven expression models were capturing generalizable genotype-phenotype relationships. After model fitting in the UKBB, the parameters estimated in this dataset ($\hat{\delta}$ and$\hat{\pi}$; $\hat{\theta}$ were independently re-estimated to account for differences in the background symptom frequency distributions across biobanks) were used to compute disease expression probabilities in AoU (using Eqn. 1). For this procedure to work, the pLoF carriers in this new dataset had to share at least one symptom in common with the pLoF carriers in the UKBB. Due to differences in clinical data encodings, this eliminated 15 diseases from our analyses. Moreover, symptom overlap across the two biobanks was rarely identical. Typically, the UKBB and AoU both harbored some unique, disease-specific symptoms. To enable replication of the UKBB-derived models within AoU, we dropped any symptoms that were unique to either dataset from the expression model. For AoU, this was straightforward, as the unique symptoms could simply be eliminated from the analysis. For the symptoms unique to the UKBB, these were removed from the disease-specific expression models (denoted $P\left( \vec{S}_{i} | \hat{\delta} \right)$) by integrating the probability distributions over the sets that contained unique symptom(s):

$$P\left( \vec{S}_{i} \setminus S_{j} | \delta\right)=\sum_{m=1}^{M_{\text{Obs}}+1} \delta_{m}\times\boldsymbol{1(}\vec{s}\equiv\mathcal{[S}_{\mathcal{m}}\setminus S_{j}]),$$

where $S_{j}$ denotes the symptom(s) to be removed from the analysis. This process effectively “aligned” the UKBB disease-specific expression models to the symptom data contained within AoU. After performing model alignment (conducted automatically by the LoadPenetranceModel function in the SymptomSetModel package^2^), disease expression in the *i*th pLoF carrier was estimated using Eqn. 1 (implemented by the PredictExpression function).

Note, the model alignment procedure almost certainly led to a loss of penetrance information in the AoU dataset, as symptoms dropped from the analysis could have been used to identify expressed pLoFs. Therefore, this could represent one of the factors that lead to reduced pLoF penetrance in this biobank. However, we doubt that it had a significant impact on our analyses for several reasons. First, disease-specific pLoF penetrance estimates were systematically higher in the AoU dataset, suggesting that the overall impact of this alignment procedure on penetrance estimates was minimal. Second, symptom-driven expression model performance on a simple validation task (see below) was similar in the UKBB and AoU. Finally, even if a substantial bias were present, it could not explain the primary finding of our machine learning analysis, as variant-specific genomic features should not correlate with missing clinical data.

*Validating the Symptom-Driven Expression Estimates*

Computing the symptom-driven disease expression probabilities was relatively straightforward. Validating their performance, however, was more challenging, as no “gold-standard” disease expression dataset exists. In other words, the previously described model yielded expression probabilities for every eligible pLoF carrier in both biobanks. However, it did not ensure that these probabilities were clinically meaningful. For example, if pLoF carrier *A* has an expression probability of 0.99 and carrier *B* has an expression probability of 0.5, it’s difficult to prove that these probabilities are capturing a coherent trend in symptom severity. To provide evidence that was true, we turned to the set of diseases that have both diagnostic *and* symptom data available in the biobanks. Because we excluded diagnoses from the symptom sets annotated to each disease, these observations could be used to validate the symptom-driven measurements.

More specifically, if our symptom-driven model were effective at capturing disease expression, then the symptom-derived probabilities should be predictive of disease diagnoses, as subjects with greater symptom burden should be more likely to receive a target diagnosis. Thus, predicting disease diagnoses using the symptom-derived probabilities represents a simple way to validate the model. After identifying diseases with both symptom-driven expression and diagnostic measurements in both the UKBB and AoU (*N*=25), we used the symptom-driven measurements to predict diagnoses, assessing performance using precision-recall curves (Supplementary Figure 2B). In these curves, each point represents a unique symptom-driven expression probability threshold. For each, we identified the pLoF carriers with symptom-driven expression probabilities exceeding this threshold. We then computed the fraction of pLoF carriers within this set that harbored the target disease diagnoses (precision, y-axis), comparing this to the total fraction of diagnosed pLoF carriers recovered by the subset (recall, x-axis). A perfect model would recover 100% of the diagnosed carriers with perfect precision (red-dotted lines). A random model would perform no better than the baseline diagnostic rates (gray dotted lines).

Clearly, the symptom-driven expression measurements predicted disease diagnoses significantly better than random (13-fold and 7.5-fold better in the UKBB and AoU, respectively; bootstrapped *P*-values < 0.0001, see Supplementary Figure 2B). Moreover, there was not a substantial difference in model performance between the UKBB and AoU, although performance was certainly better in the UKBB (likely due to training bias in the UKBB, see above). This suggests that symptom-driven expression probabilities are predictive of disease expression, at least according to diagnoses. However, it’s important to note that there are many pLoF carriers with diagnoses (approximately 40% in both biobanks) that completely lack symptom expression. Similarly, even subjects with very high symptom-driven expression scores (>0.99) sometimes lacked diagnoses. Therefore, there were clearly disagreements between symptom-driven and diagnosis-driven expression estimates.

To determine if these disagreements were statistically significant, we identified the symptom-driven expression probabilities that maximized the harmonic mean of the precision and recall scores for diagnosis prediction (also known as the F_1_-measure; red stars in Supplementary Figure 2B; equivalent to $P\left( E_{i}=1 | \vec{S}_{i},\hat{\delta},\hat{\theta},\hat{\pi} \right) \approx0.97$in both datasets). We then binarized the symptom-driven expression probabilities according to this threshold, setting any estimate to 1 if it exceeded this threshold and 0 otherwise. This binarization procedure allowed us to compute the rate of disagreement between the symptom-driven and diagnosis-driven expression measurements (i.e. number of subjects with symptom-driven expression but no diagnosis and vice versa). Although disagreements occurred (2.7% of the time in the UKBB and 4.5% in AoU), their frequency was not statistically significant in either biobank (McNemar’s Test *P­*-values = 0.16 and 0.12 in the UKBB and AoU respectively).

*The Variant-Intrinsic Features Used for pLoF Penetrance Prediction*

This section describes the variant-intrinsic genomic features that were used to build the phenotype expression prediction models for each pLoF class. The feature descriptions are separated by whether they were used in all three prediction models (variant-type agnostic) or were specific to a particular class. We hypothesize that these features correlate with “leaky” or “incomplete” loss-of-function, although additional functional studies will be needed to confirm this. Note, these features were mostly derived from prior analyses focused on pLoF annotation interpretation in biobanks^3–5^.

Variant Type Agnostic Features:

- CADD Score^6^: The Combined Annotation-Dependent Depletion (CADD) score predicts the deleteriousness of individual variants using a single numerical score derived from a wide range of variant-specific features, including but not limited to evolutionary conservation, DNA sequence motifs, and predicted impact on biochemical activity. Uniquely, CADD does not build these scores by training on a set of variants known to cause human disease. Instead, the scores are inferred by fitting a machine learning model to a set of evolutionarily neutral variants (proxy-negative cases) and a set of simulated mutations, which may or may not be deleterious (proxy-positive cases). This makes CADD well-suited for the analyses conducted in this study, as the score should not be polluted with information from prior ClinVar annotations.
- LOFTEE Confidence Flag^7^: The LOFTEE plug-in for VEP^8^ not only identifies putative loss-of-function variants but also assigns them a confidence flag (low or high) based on several variant-specific features (e.g. distance from end of transcript, ancestral alleles, etc.; see [https://github.com/konradjk/LOFTEE](https://github.com/konradjk/loftee) for details)
- Transcript Type^9^: All variants were assigned to one of three transcript types (MANE Select, MANE Plus Clinical, Other) based on the most clinically relevant transcript that was predicted to be impacted by the annotation software.
- Relative Amino Acid Position^5^: This feature captures the relative position of a variant within the amino acid sequence. It is defined slightly differently for the three variant classes. For stop gain variants, this feature is equivalent to the fraction of the amino acid sequence predicted to be lost if a variant escapes NMD escape. Thus, the feature simply captures a pLoF’s relative distance from the N-terminus of protein (according to the MANE Select^9^ transcript). For frameshift variants, this feature computes the relative fraction of amino acids predicted to be altered by an expressed frameshift, assuming it escapes non-sense mediated decay. For splice change variants, determining the exact location of the last normal amino acid can be challenging. Therefore, we set the last normal amino acid to be the residue just proximal to the impacted splice site in the transcript model. This could clearly be improved (ex: by considering in-frame splice rescue events, exon skipping, etc) but will be the focus of future work.

Stop-Gain Variant Features:

- Predicted Non-sense Mediated Decay (NMD) Escape^10^: It is well known that some stop gain variants escape non-sense mediated decay, enabling the expression of a potentially functional but truncated transcript. To predict NMD escape, we used the decision tree developed in Lindeboom et al^11^. Note, we did not encode predicted NMD Escape using a binary annotation (Present, Absent) but instead included the reason for the predicted escape into the model (No NMD Escape Present, Last Exon, First Exon ≤ 150nt from Start, Large Exon, ≤50nt from Last Exon-Exon Junction).
- Possible Methionine Rescue (Translation Re-initiation)^3^: If a stop-gain variant occurs early enough in the amino acid sequence, then translation can potentially be rescued by another methionine residue that occurs just downstream. The exact criteria needed to be met for this to occur are unknown and may be variable across proteins. For this analysis, a stop gain variant had to meet the following criteria to flag for possible methionine rescue: 1) located in the first exon and 2) have a downstream methionine for alternate translation initiation that truncated <10% of the total protein length.

Frameshift Variant Features

- Last Coding Exon^3^: This is a simple binary feature that indicates if the frameshift occurred in the last exon.
- Possible Methionine Rescue^3^: This feature is computed in the same fashion for frameshift and stop gain variants.
- Note, NMD escape is certainly possible for frameshift variants with the added complexity that the escape is occurring on a frameshifted sequence. It’s possible that additional features based on NMD escape would improve frameshift penetrance prediction, but additional work is needed to determine when these rules may apply.

Splice Change Variant Features

- SpliceAI Score^12^: SpliceAI is a deep learning model that predicts changes in the splicing probabilities at different sites induced by a genetic variant relative to the splicing probabilities for the reference sequence (assuming some specific transcript model). For the current analysis, we re-computed SpliceAI scores using the Ensembl transcripts for each gene (Release 109), allowing for a maximum of 500bp between the variant and impacted site. For expression prediction, the maximum SpliceAI score (maximum difference in splicing probability between the reference and mutated transcript) was included as a feature. Note, several additional features were derived from the SpliceAI output. These are outlined in detail below.
- Splice Mutation Type: Pathogenic splice mutations can impact transcript structure in complex ways, sometimes inducing multiple changes simultaneously. For the sake of simplicity, we used the SpliceAI output to assign each mutation to one of five classes based on the highest SpliceAI score observed for the variant: Donor Gain, Donor Loss, Acceptor Gain, Acceptor Loss and Indeterminate (i.e. maximum SpliceAI score = 0.0 or NaN).
- Outside Coding Region: Some splice sites occur in exons that lie outside the coding region. Although they could result in loss-of-function, many of these may be tolerated. Therefore, we included a binary feature that flagged splice variants predicted to impact only non-coding exons.
- Last Coding Exon^3^: This feature indicates whether a splice mutation is predicted to impact the last coding exon. Like the other variant classes, such mutations are more likely to result in residual allelic activity.
- Persistent Original Splice Site Score: Sometimes, SpliceAI predicts that the original splice site remains intact with some non-zero probability, which may be indicative of leaky wild type expression. Therefore, we computed the difference between the SpliceAI score for the original and derived sites. Generally, this is simply equivalent to the global SpliceAI score, but other times, a variant increases the splicing probability for the wildtype splice site along with the derived site. This feature accounts for this phenomenon.
- In-frame Exon Rescue^3^: If the exon impacted by a splice change has a nucleotide length that is a multiple of 3, then it can theoretically be skipped without disrupting the reading frame. This phenomenon was accounted for in the model using a binary feature (Present, Absent).
- Possible Methionine Rescue^3^: For splice variants, this is a less likely rescue mechanism. Nevertheless, given that a variant impacts the first exon, we allowed for possible methionine rescue assuming that there was a methionine residue in the second exon that truncated less than 10% of the amino acid sequence.
- In-frame Intron Retention^3^: If the intron to be spliced out has a nucleotide length that is a multiple of 3, then it can potentially be retained without impacting the transcript reading frame. This phenomenon is accounted for in the model using a binary feature (Present, Absent).
- Cryptic Rescue Score^3,4^: Many times, when SpliceAI predicts a primary splice site change, a secondary change is predicted to occur simultaneously that could negate the impact of the primary change. More specifically, if a genetic variant is predicted to cause a donor (acceptor) loss event in a transcript, there can be a complementary donor (acceptor) gain event just upstream/downstream of the predicted loss site but with a lower SpliceAI score. If this event remains in-frame with the original transcript, then the impact of the mutation may be minimal, as this complementary site could compensate for the loss. Alternatively, many donor (acceptor) gain events occur in-frame with the original donor (acceptor) site. So as long the downstream acceptor (upstream donor) site remains intact, then the impact of the variant may be insignificant. This Cryptic Rescue Score summarizes both possible rescue events using the output from SpliceAI. For primary splice site loss events (donor or acceptor), the Cryptic Rescue Score is simply the SpliceAI score for the in-frame gain event (assigned 0.0 if no in-frame gain is predicted). For primary in-frame gain events (donor or acceptor), the Cryptic Splice Score is harder to define. For this analysis, we used the corresponding splice site loss score (ex: a loss score of 1.0 should indicate that this predicted in-frame gain is preferentially being used) but acknowledge that this very much imperfectly captures the phenomenon. Clearly, more work is needed to effectively capture the complexity of splice mutation rescue events.

*Training pLoF Penetrance Prediction Models in the UKBB*

In the main text, we defined the following probabilistic model for pLoF penetrance prediction:

$$\begin{aligned} \mathcal{P}_{i}\mathcal{=F}\left( \vec{V}_{i};\theta\right), \end{aligned}$$

where $\mathcal{P}_{i}$ is the disease expression measurement in the *i*th subject (i.e. $\mathcal{P}_{i}$ = $\text{max}\left[ D_{i},P\left( E_{i}=1 | \vec{S}_{i},\hat{\delta},\hat{\theta},\hat{\pi} \right) \right]$) and $\mathcal{F}$ is some function that maps the vector $\vec{V}_{i}$onto disease expression probability space (via a parameter set $\theta$). Typically, these models are trained on binary outcome data (i.e. $\mathcal{P}_{i}\in0,1$). However, our symptom-driven expression estimates were continuous probabilities within the interval of 0 and 1 (i.e. $\mathcal{P}_{i}\in[0,1]$), complicating training. Note, this is not an issue for validation, as the statistic we used to assess model performance (average penetrance score) does not require binary outcome data.

To facilitate model training in the UKBB, we binarized the symptom-driven expression probabilities by selecting a single threshold for disease expression. Those pLoF carriers with expression probabilities above this threshold were deemed to express the disease of interest, while those with scores below the threshold were not. More specifically, we considered a pLoF to be expressed for the purpose of model training if the carrier harbored a Mendelian disease diagnosis (i.e. $D_{i}=1$, assuming diagnostic data was available) or if their symptom-driven expression probability exceeded the F_1_ threshold depicted as a red star in Supplementary Figure 2B (i.e. $P\left( E_{i}=1 | \vec{S}_{i},\hat{\delta},\hat{\theta},\hat{\pi} \right)\geq F_{1}$, where $F_{1}=0.97$ for the UKBB). To avoid confounding the model with symptomatic pLoF carriers whose expression probabilities did not exceed the $F_{1}$ threshold, we removed any (weakly) symptomatic carrier whose binarized disease expression score was equal to zero (i.e. $0<P\left( E_{i}=1 | \vec{S}_{i},\hat{\delta},\hat{\theta},\hat{\pi} \right) < F_{1}$). Following binarization and filtering, we were left with 9,498 UKBB pLoF carriers who were clearly symptomatic/asymptomatic. These pLoF carriers were exclusively used for penetrance prediction model training.

Model training in the UKBB was performed using 5-fold leave-one-out cross validation, and performance on the withheld subset was assessed using the increase in the average penetrance score (see main text) from baseline. In Supplementary Figure 4, the performance of the random forest prediction model in cross-validation experiments is compared to a simpler penalized logistic regression model. The random forest model had relatively consistent performance across the cross-validation samples and consistently outperformed logistic regression. Therefore, this model was used to produce all the machine learning results presented in the main text.

*Penetrance Prediction Model Feature Importance Analysis*

To broadly capture incomplete or “leaky” loss-of-function, a diverse set of features were included into each of our penetrance prediction models (see above). However, they were not *a priori* guaranteed to be predictive of penetrance. Moreover, many of these features were highly correlated, and therefore, likely capture redundant information. As a result, the contribution of each feature to penetrance prediction was largely unknown, even after demonstrating that the models were generally predictive of pLoF penetrance across a diverse population of carriers (see Main Test Figure 5).

To investigate the contribution of each variant-specific feature to penetrance prediction, we computed their carrier-specific Shapley additive explanation (or SHAP) values^13^ within the AoU validation cohort. Briefly, SHAP values use principles derived from cooperative game theory to quantify the additive contribution of each feature to the predictions made by a machine learning model (see Ponce-Bobadilla et al.^14^ for a biologically-relevant tutorial). These statistics have several desirable properties (ex: the sum of the feature-specific SHAP values for each observation is equivalent to the total prediction made by model), and they can be computed exactly in some cases^15^ but are generally approximated using dataset sampling^16^. For this analysis, we computed the SHAP values for the features included into our random forest penetrance prediction models using the TreeExplainer function available in the shap^17^ package. Rather than computing exact SHAP values on the training dataset, we approximated feature importance in the AoU validation dataset using random sampling (using the **‘feature_perturbation = interventional’ argument for the** TreeExplainer function**).**

**The results of this analysis for each of the three pLoF prediction models are displayed in Supplementary Figures 5-7. Overall, CADD scores**^6^ **were consistently among the most predictive features, likely because they summarize multiple forms of genomic evidence that capture variant “deleteriousness.” That said, variant-class specific features often contributed strongly to the penetrance predictions. For example, in the case of stop gain variants, having no features that predicted NMD escape consistently resulted in higher penetrance scores. Moreover, for splice change variants, statistics derived from a probabilistic splicing model**^12^ **(Splice AI Score, Cryptic Rescue Score) contributed strongly to the penetrance estimates. Additional results for each variant class prediction model are further discussed in the legends for Supplementary Figures 5-7.**

**Supplementary Figures and Tables**

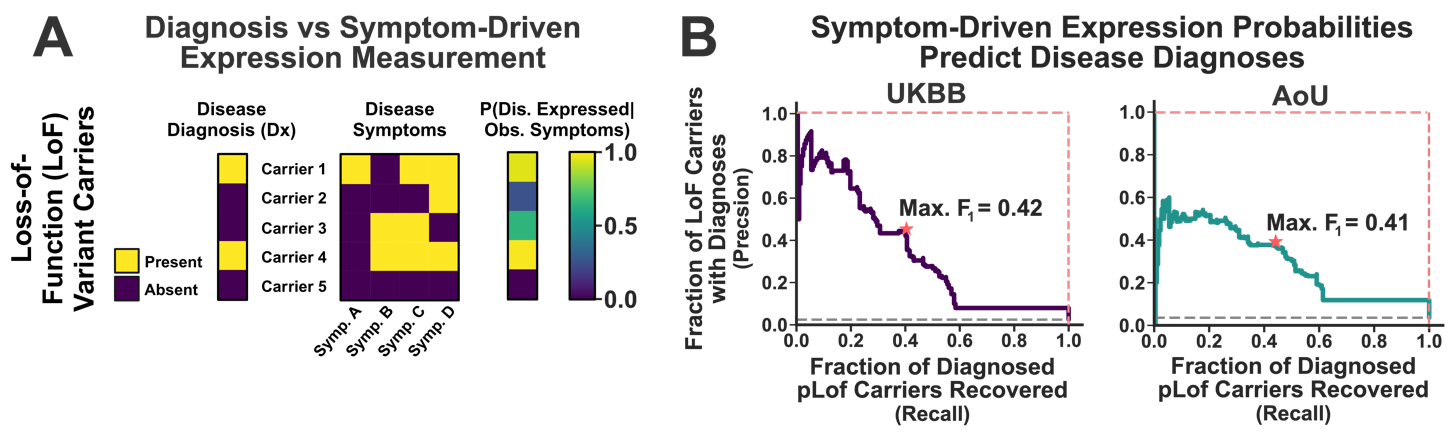

Supplementary Figure 2: *Estimating and Validating Symptom-Driven Disease Expression*. (A): A simple illustration of the symptom-driven expression model. (B): Symptom-driven expression probabilities predict rare disease diagnoses in both datasets. These panels display the precision (i.e. fraction of LoF carriers with diagnoses; y-axis) and the recall (i.e. total fraction of diagnosed pLoF carriers recovered; x-axis) for a model that uses symptom-driven expression probabilities to identify pLoF carriers with disease diagnoses. Each point on these curves represents a different expression probability threshold for identifying carriers at-risk for diagnosis. The red stars denote the expression score (≈0.97 in both datasets) that maximized the F_1_-measure (harmonic mean of precision and recall) for the predictions. Gray lines indicate the performance of a random model; red-dashed lines the performance of a perfect classifier. The left panel depicts performance in the UKBB (training dataset; *N* = 8,242 pLoF carriers), while the right panel depicts performance in AoU (validation dataset; *N* = 6,129 pLoF carriers). Created using Biorender.com.

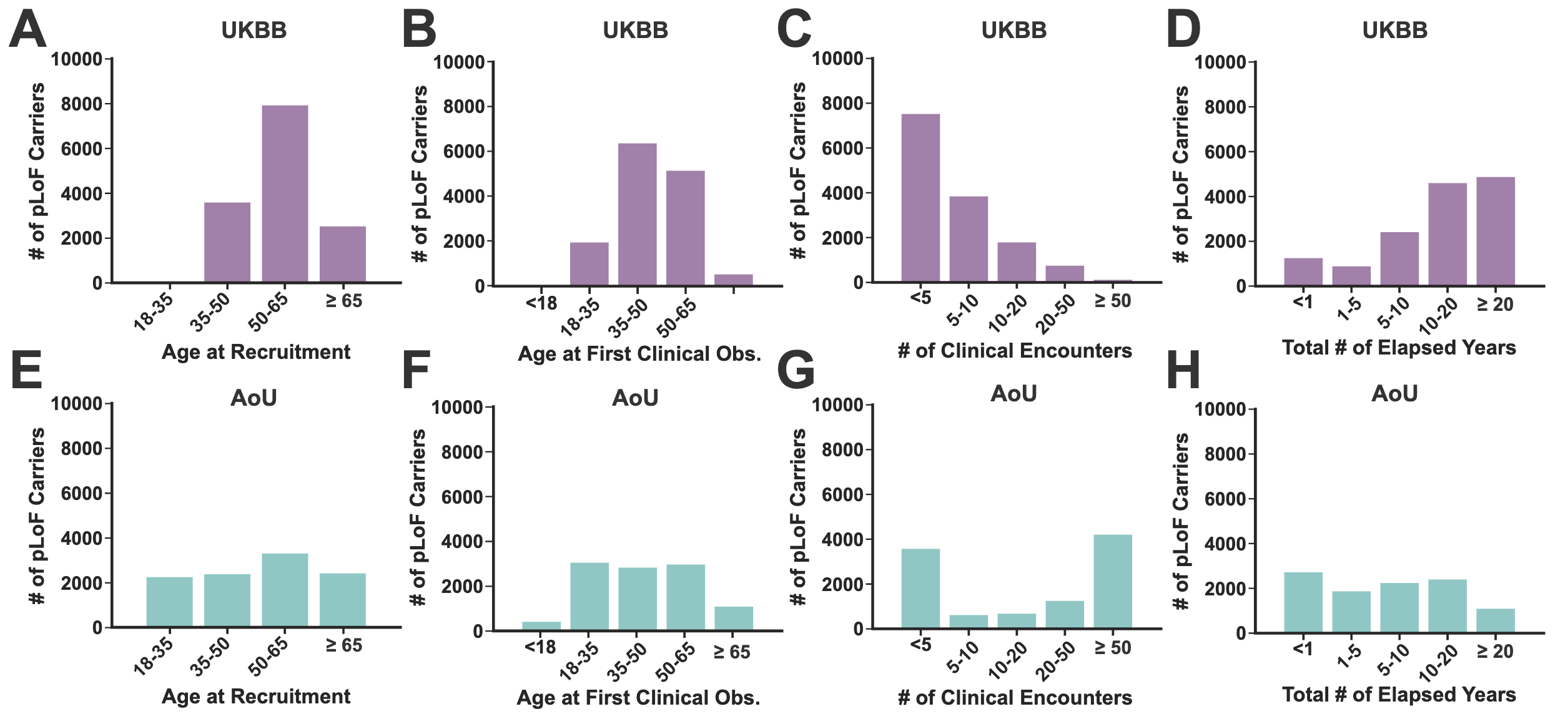

Supplementary Figure 1: Clinical data coverage distributions among pLoF carriers in the UKBB and AoU. (A, E): Distribution of age at the time of recruitment (A: UKBB; E: AoU). (B, F): Distribution of age at the first clinical observation in the EHR (B: UKBB; F: AoU). (C, G): Distribution of total number of clinical visits in the EHR (C: UKBB; G: AoU). (D, H): Distribution of total number of elapsed years between the first and last clinical visits (D: UKBB; H: AoU).

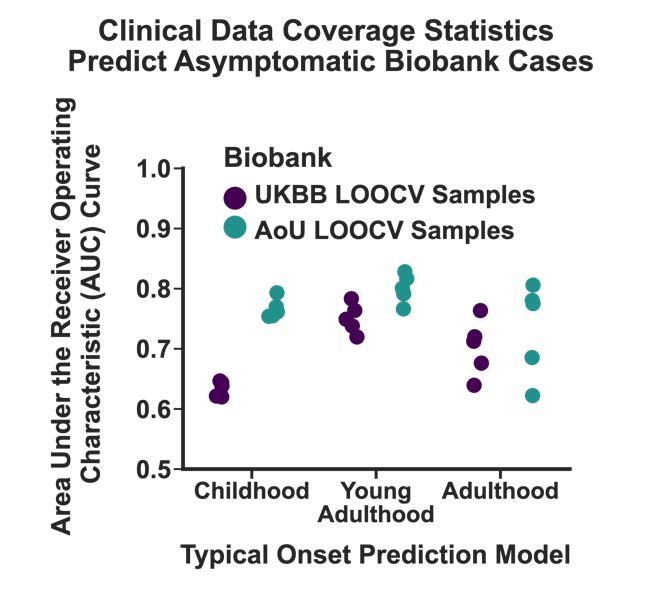

Supplementary Figure 3: Predicting Asymptomatic pLoF Carriers using Clinical Data Coverage. The coverage statistics displayed in Supplementary Figure 1 were used to build models to predict whether the pLoF carriers (stratified by disease onset; x-axis) would be asymptomatic for their target phenotypes (disease diagnoses and/or symptoms). Model performance was assessed using the area under the receiver operating characteristic curve (AUC; y-axis) using leave-one-out 5-fold cross validation (LOOCV). All models consistently performed better than random (AUC=0.5), with some variability in performance both within and between the different onset classes and biobanks. For the most part, asymptomatic status was easiest to predict for the young adult-onset disorders. We suspect that multiple factors contribute to this, including the more uniform clinical data coverage for this age interval (Supplementary Figure 1) coupled with the fact that pLoF carriers with severe, childhood-onset diseases may be depleted from these biobanks. This is particularly true for the UKBB, where asymptomatic status for childhood-onset diseases was the most difficult to predict (left). This likely stems from this dataset’s recruitment strategy, which focused on healthy adults and may therefore lead to the depletion of subjects that are symptomatic from childhood-onset diseases.

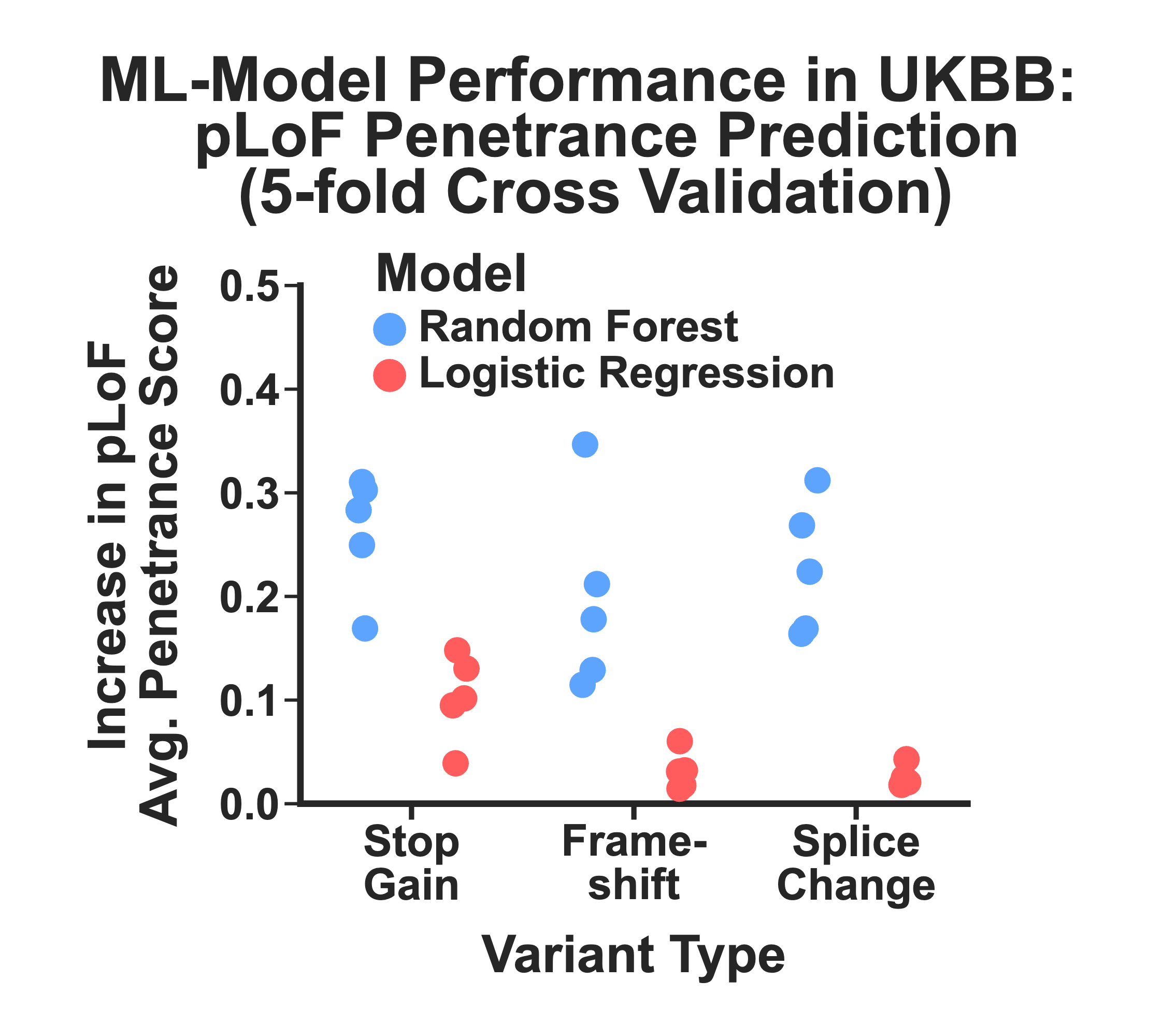

Supplementary Figure 4: pLoF penetrance prediction model training results (UKBB). We evaluated the performance of two different machine learning frameworks for predicting pLoF penetrance using genomic features: Logistic Regression (red) and Random Forests (blue). We assessed performance using the average increase in penetrance achieved by filtering pLoFs in the UKBB according to their expression scores (see Main Text for details) using leave-one-out 5-fold cross validation. Random forest models consistently outperformed logistic regression in these experiments for all three variant classes considered in this study.

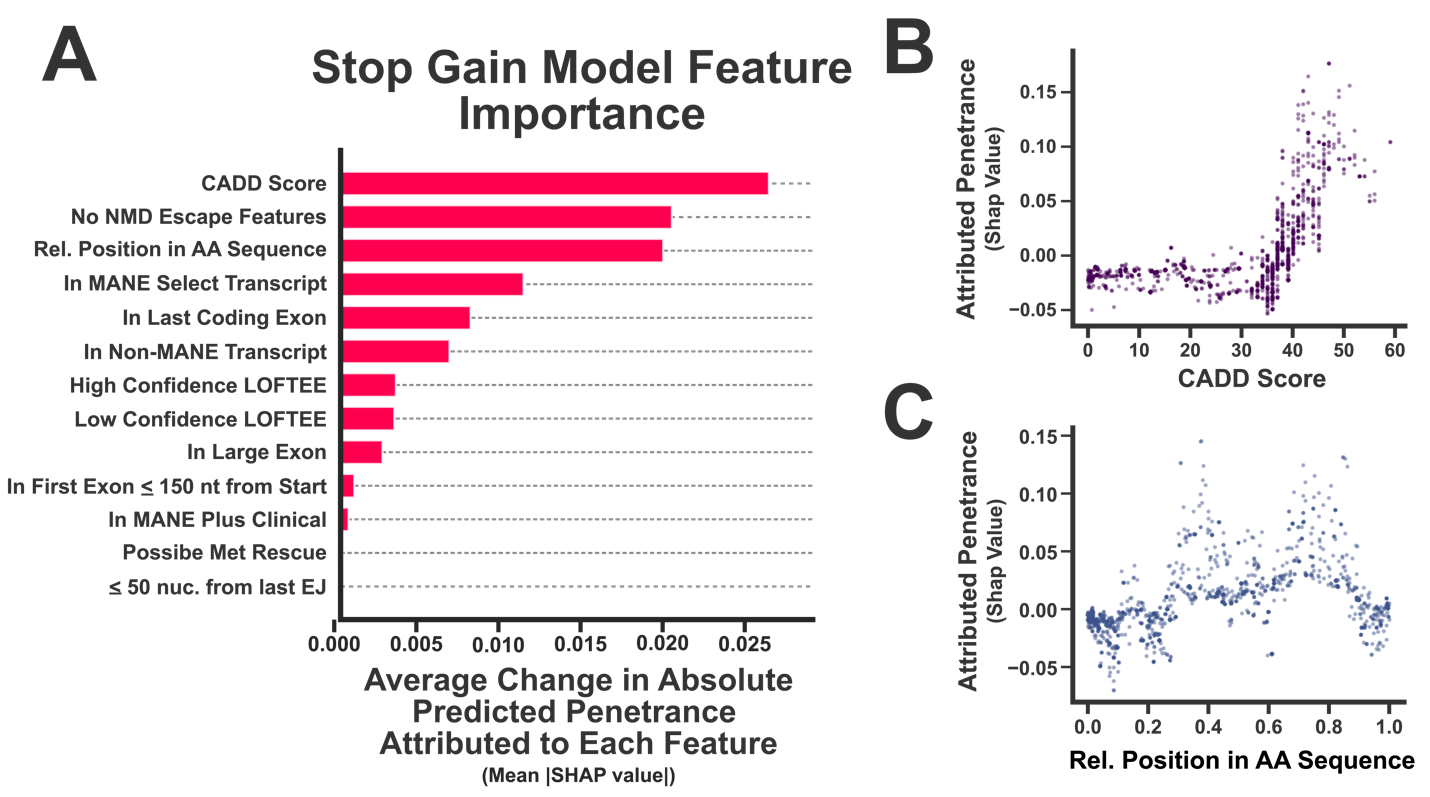

*Supplementary Figure 5: Feature importance analysis for the stop gain penetrance prediction model.* (A): The importance of each feature to the model’s penetrance predictions was quantified using the change in absolute predicted penetrance attributed to each feature, averaged across the stop gain carriers in the AoU validation dataset (equivalent to the mean of the absolute value of the SHAP statistics for this feature). (B): The penetrance attributed to the CADD score is plotted against its numerical value for each stop gain carrier in the AoU validation dataset. (C): Same as in (B), except the penetrance attributed to the relative position in the amino acid sequence is displayed instead. As expected, attributed penetrance follows a monotonically increasing relationship for CADD scores. However, the relationship between relative amino acid position and penetrance is far more complex.

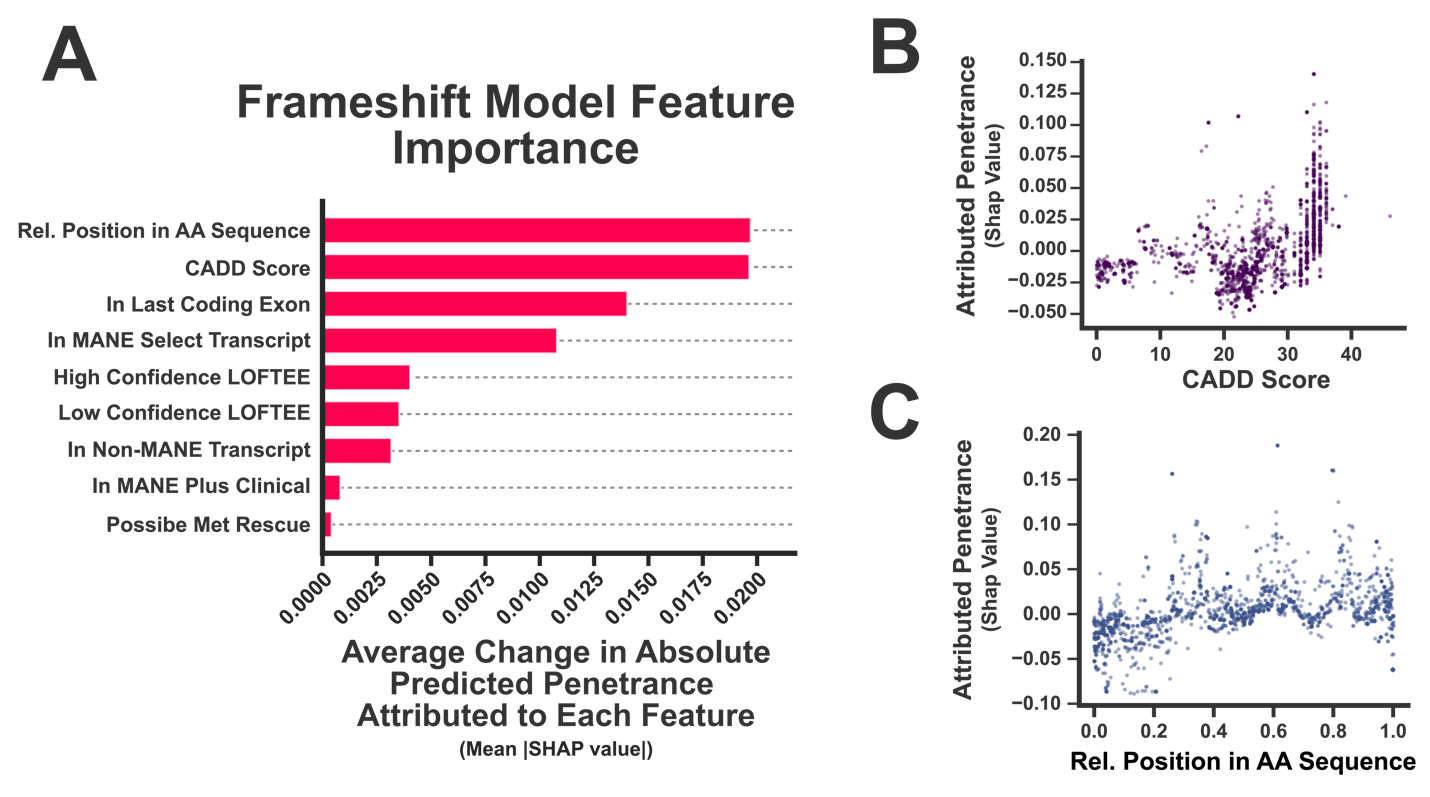

*Supplementary Figure 6: Feature importance analysis for the frameshift penetrance prediction model.* (A): The importance of each feature to the model’s penetrance predictions was quantified using the change in absolute predicted penetrance attributed to each feature, averaged across the frameshift carriers in the AoU validation dataset (equivalent to the mean of the absolute value of the SHAP statistics for this feature). (B): The penetrance attributed to the CADD score is plotted against its numerical value for each frameshift variant carrier in the AoU validation dataset. (C): Same as in (B), except the penetrance attributed to the relative position in the amino acid sequence is displayed instead. For frameshift variants, attributed penetrance again follows a monotonically increasing relationship for CADD scores, although the relationship is more variable when compared to stop gain variants. The relationship between relative amino acid position and frameshift variant penetrance is again complex.

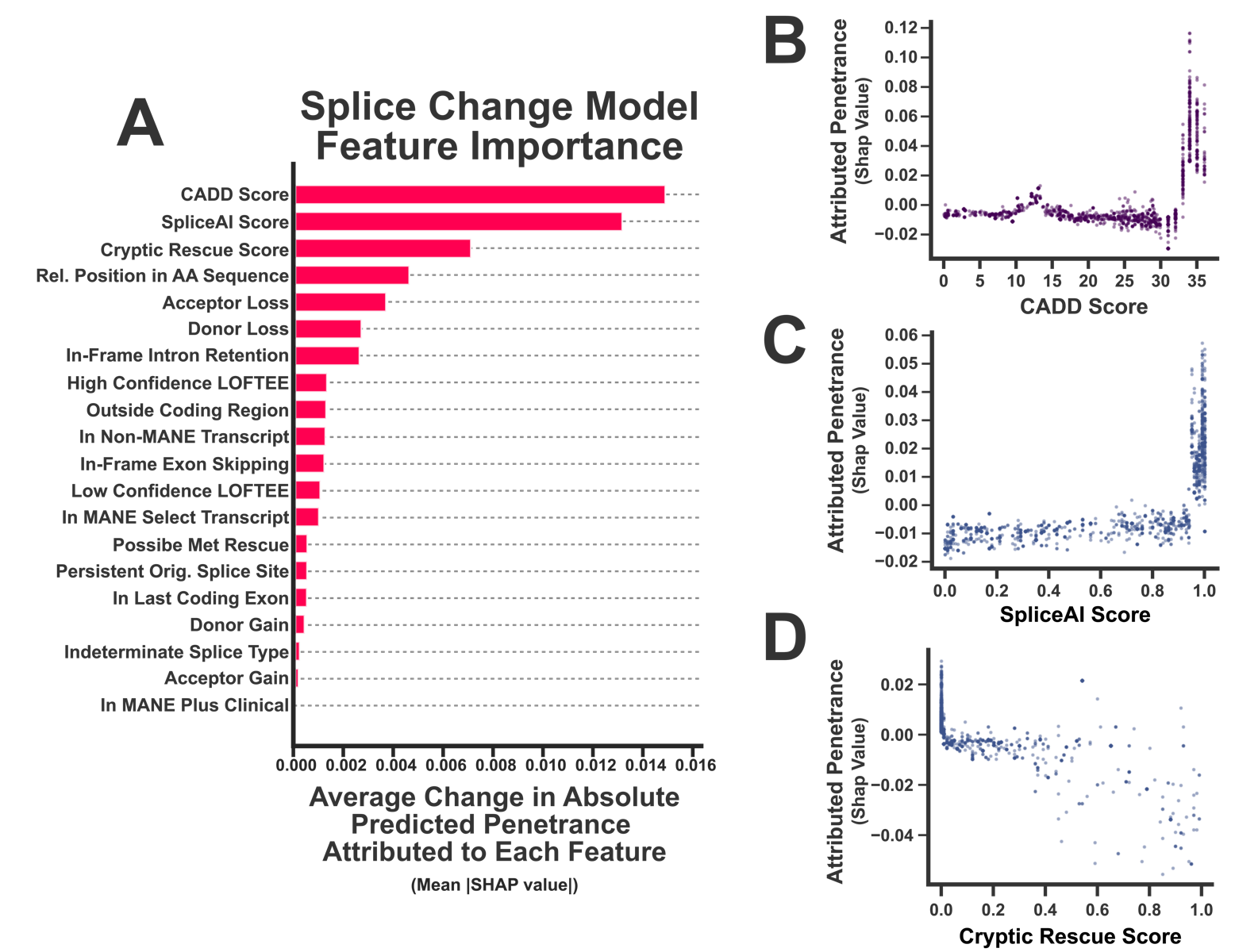

*Supplementary Figure 6: Feature importance analysis for the splice change penetrance prediction model.* (A): The importance of each feature to the model’s penetrance predictions was quantified using the change in absolute predicted penetrance attributed to each feature, averaged across the splice change carriers in the AoU validation dataset (equivalent to the mean of the absolute value of the SHAP statistics for this feature). (B): The penetrance attributed to the CADD score is plotted against its numerical value for each splice change variant carrier in the AoU validation dataset. (C): Same as in (B), except the penetrance attributed to the SpliceAI score is displayed instead. (D): Same as in (B), except the penetrance attributed to the Cryptic Rescue Score is depicted. For splice change variants, attributed penetrance follows a strong, monotonic relationship with CADD scores, which is mirrored in the SpliceAI scores (likely because splicing prediction scores have been incorporated into CADD). As expected, low Cryptic Rescue Scores are associated with increased penetrance. However, moderate-to-high scores are not necessarily indicative of low penetrance, suggesting that much remains to be learned about the complexities of splice disruption rescue events.

|  | **Average Penetrance Score**  **(95% Confidence Interval)** | | |
| --- | --- | --- | --- |
| **pLoF Filtering**  **Method** | **All Variants in AoU** | **Variants Unique to AoU** | **Variants**  **Shared with UKBB** |
| Baseline Penetrance | 10.4% | 13.6% | 7.1% |
| MANE Select+High-Confidence LOFTEE Flag | 12.1%  (11.2%-13.0%) | 16.1%  (14.7%-17.5%) | 7.0%  (6.1%-7.9%) |
| Pathogenic/Likely Pathogenic (P/LP) in ClinVar Only | 12.6%  (11.6%-13.6%) | 16.6%  (15.1%-18.2%) | 7.8%  (6.7%-9.0%) |
| Machine Learning  Model | 22.7%  (20.2%-25.6%) | 21.3%  (19.0%-24.4%) | 27.7%  (22.2%-33.8%) |
| Machine Learning  Model, P/LP in ClinVar Only | 34.8%  (30.0%-39.7%) | 28.8%  (24.8%-34.0%) | 52.9%  (42.9%-62.0%) |

Supplementary Table 1: The impact of shared variants on pLoF penetrance prediction in AoU. This table compares different methods for risk stratifying pLoFs by penetrance. The first row provides the baseline penetrance for a particular variant set. The second and third rows represent simple binary filters (see Main Text for details), while the latter two rows filter variants using scores derived from our machine learning penetrance prediction models. The cells display the average pLoF penetrance score for each filter (along with a bootstrapped 95% confidence interval). These estimates were obtained by summing the average pLoF penetrance across all possible scoring thresholds, weighting them by the loss of total penetrance (i.e. recall) incurred at each step (i.e. average penetrance score^18^, see Main Text Methods for details). The columns compare the performance of these methods on all variants (left), those that are unique to AoU (i.e. variants absent from the UKBB; middle), and those that are shared by both biobanks. The penetrance prediction models do perform worse on the unique versus the shared variants, which is not unexpected. However, this does not change the general conclusions of our analysis. Moreover, it highlights the replicability of the genotype-phenotype measurements across biobanks. Finally, the machine learning model-derived penetrance prediction models significantly outperform the simple filters for all three sets of variants (bootstrapped P-values < 1.0×10^-4^ for all comparisons).

**References**

1.	Jordan MI, Ghahramani Z, Jaakkola TS, Saul LK. An Introduction to Variational Methods for Graphical Models. Machine Learning. 1999 Nov 1;37(2):183–233.

2.	daverblair/SymptomSetModel: A symptom-driven probability model for predicting disease expression [Internet]. [cited 2024 Dec 3]. Available from: https://github.com/daverblair/SymptomSetModel

3.	Singer-Berk M, Gudmundsson S, Baxter S, Seaby EG, England E, Wood JC, Son RG, Watts NA, Karczewski KJ, Harrison SM, MacArthur DG, Rehm HL, O’Donnell-Luria A. Advanced variant classification framework reduces the false positive rate of predicted loss-of-function variants in population sequencing data. Am J Hum Genet. 2023 Sep 7;110(9):1496–1508. PMCID: PMC10502856

4.	de Sainte Agathe JM, Filser M, Isidor B, Besnard T, Gueguen P, Perrin A, Van Goethem C, Verebi C, Masingue M, Rendu J, Cossée M, Bergougnoux A, Frobert L, Buratti J, Lejeune É, Le Guern É, Pasquier F, Clot F, Kalatzis V, Roux AF, Cogné B, Baux D. SpliceAI-visual: a free online tool to improve SpliceAI splicing variant interpretation. Human Genomics. 2023 Feb 10;17(1):7. PMCID: PMC9912651

5.	Beaumont RN, Hawkes G, Gunning AC, Wright CF. Clustering of predicted loss-of-function variants in genes linked with monogenic disease can explain incomplete penetrance. Genome Medicine. 2024 Apr 26;16(1):64. PMCID: PMC11046769

6.	Schubach M, Maass T, Nazaretyan L, Röner S, Kircher M. CADD v1.7: using protein language models, regulatory CNNs and other nucleotide-level scores to improve genome-wide variant predictions. Nucleic Acids Research. 2024 Jan 5;52(D1):D1143–D1154. PMCID: PMC10767851

7.	Karczewski KJ, Francioli LC, Tiao G, Cummings BB, Alföldi J, Wang Q, Collins RL, Laricchia KM, Ganna A, Birnbaum DP, Gauthier LD, Brand H, Solomonson M, Watts NA, Rhodes D, Singer-Berk M, England EM, Seaby EG, Kosmicki JA, Walters RK, Tashman K, Farjoun Y, Banks E, Poterba T, Wang A, Seed C, Whiffin N, Chong JX, Samocha KE, Pierce-Hoffman E, Zappala Z, O’Donnell-Luria AH, Minikel EV, Weisburd B, Lek M, Ware JS, Vittal C, Armean IM, Bergelson L, Cibulskis K, Connolly KM, Covarrubias M, Donnelly S, Ferriera S, Gabriel S, Gentry J, Gupta N, Jeandet T, Kaplan D, Llanwarne C, Munshi R, Novod S, Petrillo N, Roazen D, Ruano-Rubio V, Saltzman A, Schleicher M, Soto J, Tibbetts K, Tolonen C, Wade G, Talkowski ME, Neale BM, Daly MJ, MacArthur DG. The mutational constraint spectrum quantified from variation in 141,456 humans. Nature. 2020 May;581(7809):434–443. PMCID: PMC7334197

8.	McLaren W, Gil L, Hunt SE, Riat HS, Ritchie GRS, Thormann A, Flicek P, Cunningham F. The Ensembl Variant Effect Predictor. Genome Biology. 2016 Jun 6;17(1):122. PMCID: PMC4893825

9.	Morales J, Pujar S, Loveland JE, Astashyn A, Bennett R, Berry A, Cox E, Davidson C, Ermolaeva O, Farrell CM, Fatima R, Gil L, Goldfarb T, Gonzalez JM, Haddad D, Hardy M, Hunt T, Jackson J, Joardar VS, Kay M, Kodali VK, McGarvey KM, McMahon A, Mudge JM, Murphy DN, Murphy MR, Rajput B, Rangwala SH, Riddick LD, Thibaud-Nissen F, Threadgold G, Vatsan AR, Wallin C, Webb D, Flicek P, Birney E, Pruitt KD, Frankish A, Cunningham F, Murphy TD. A joint NCBI and EMBL-EBI transcript set for clinical genomics and research. Nature. 2022 Apr;604(7905):310–315. PMCID: PMC9007741

10.	Dyle MC, Kolakada D, Cortazar MA, Jagannathan S. How to get away with nonsense: Mechanisms and consequences of escape from nonsense-mediated RNA decay. Wiley Interdiscip Rev RNA. 2020 Jan;11(1):e1560. PMCID: PMC10685860

11.	Lindeboom RGH, Vermeulen M, Lehner B, Supek F. The impact of nonsense-mediated mRNA decay on genetic disease, gene editing and cancer immunotherapy. Nat Genet. 2019 Nov;51(11):1645–1651. PMCID: PMC6858879

12.	Jaganathan K, Kyriazopoulou Panagiotopoulou S, McRae JF, Darbandi SF, Knowles D, Li YI, Kosmicki JA, Arbelaez J, Cui W, Schwartz GB, Chow ED, Kanterakis E, Gao H, Kia A, Batzoglou S, Sanders SJ, Farh KKH. Predicting Splicing from Primary Sequence with Deep Learning. Cell. 2019 Jan 24;176(3):535-548.e24. PMID: 30661751

13.	Lundberg SM, Lee SI. A Unified Approach to Interpreting Model Predictions. Advances in Neural Information Processing Systems [Internet]. Curran Associates, Inc.; 2017 [cited 2025 Jan 31]. Available from: https://papers.nips.cc/paper_files/paper/2017/hash/8a20a8621978632d76c43dfd28b67767-Abstract.html

14.	Ponce‐Bobadilla AV, Schmitt V, Maier CS, Mensing S, Stodtmann S. Practical guide to SHAP analysis: Explaining supervised machine learning model predictions in drug development. Clin Transl Sci. 2024 Oct 28;17(11):e70056. PMCID: PMC11513550

15.	Lundberg SM, Erion G, Chen H, DeGrave A, Prutkin JM, Nair B, Katz R, Himmelfarb J, Bansal N, Lee SI. From local explanations to global understanding with explainable AI for trees. Nat Mach Intell. Nature Publishing Group; 2020 Jan;2(1):56–67.

16.	Janzing D, Minorics L, Bloebaum P. Feature relevance quantification in explainable AI: A causal problem. Proceedings of the Twenty Third International Conference on Artificial Intelligence and Statistics [Internet]. PMLR; 2020 [cited 2025 Feb 13]. p. 2907–2916. Available from: https://proceedings.mlr.press/v108/janzing20a.html

17.	Welcome to the SHAP documentation — SHAP latest documentation [Internet]. [cited 2025 Feb 13]. Available from: https://shap.readthedocs.io/en/latest/index.html

18.	sklearn.metrics.average_precision_score — scikit-learn 0.23.1 documentation [Internet]. [cited 2020 Jul 2]. Available from: https://scikit-learn.org/stable/modules/generated/sklearn.metrics.average_precision_score.html
